# PortalEvent: An Event-Based Ontology for Patient-Initiated Portal Communication

**DOI:** 10.64898/2026.06.01.26354623

**Authors:** Jialin Yang, Joseph Gatto, Roshnik Rahat, Parker Seegmiller, Timothy E. Burdick, Martin Keil, Stephen K. Liu, Sarah M. Preum

**Author notes:** Corresponding Author: Jialin Yang, Department of Computer Science, Dartmouth College, Address: 15 Thayer Drive, Hanover, NH 03755, United States, Sarah M. Preum, Department of Computer Science, Dartmouth College. These authors contributed equally (Jialin Yang, Joseph Gatto).

## Abstract

**Background:** Patient portal messaging has become a primary channel for asynchronous clinical communication. The messages cover a wide range of topics, from symptom reporting and medication concerns to administrative requests. In recent years, message volume has grown substantially. When a message lacks information the clinician needs, it often leads to back-and-forth that delays resolution of the patient’s request, and it also takes time from clinicians that could be spent on more urgent matters. To build automated tools that reduce the burden, a structured representation is needed to understand what details these messages contain or lack, so the automated tool can provide help accordingly. However, there is no existing scalable, reproducible method to characterize these messages in a structured way with fine granularity.

**Objective:** To create a scalable, reproducible representation for patient portal communication, we developed an ontology schema grounded in real portal messages and extrinsically validated it by characterizing patient portal messages and clinician follow-up questions in a large electronic health record (EHR) portal dataset from a real health system.

**Methods:** We developed the ontology through three stages: Literature review, large language model (LLM)–assisted induction and iterative expert review in collaboration with clinical natural language processing (NLP) researchers and three clinicians. We validated the ontology on real patient messages and clinician follow-up questions, and manually verified its results on randomly selected samples.

**Results:** The final ontology defines 8 event types and 71 roles covering clinical content (eg, symptoms, medications, diagnostic tests, treatment responses, and patient context) and administrative and social content (medical needs, logistics, and social). In 10,000 randomly selected patient messages, medical needs and symptom events were the most frequent, and clinical and administrative events often co-occurred within a single message. Among 57,321 clinician follow-up questions, the most common targets were symptom, medication, and medical needs events. In manual review, average detection precision was 0.90 or higher for 6 of 8 event types, interannotator agreement on argument judgments exceeded 0.90 for every event type, and detection rates were consistent across 3 open-source LLMs.

**Conclusions:** PortalEvent is a scalable and reproducible event ontology for patient portal messages, developed and validated in collaboration with three clinicians. It can be applied at scale with open-source LLMs in a secure and training-free environment. Results show that messages often combine both clinical and administrative needs, and clinician follow-up questions concentrate on a small set of roles, which frequently seek the basic facts of an event, such as which symptom or medication is involved. These patterns may inform the design of patient- and provider-facing communication support tools.

## Introduction

Patient portal messaging has become an important channel of asynchronous communication between patients and clinicians [1], and it contributes to clinician inbox load [2,3]. Both message volume and the associated clinician workload have grown substantially in recent years [1]. Between 2020 and 2025, patients at more than 2,000 hospitals sent 1.34 billion portal messages, and clinicians and staff sent 3.25 billion [1]. As portal messaging grows, it becomes essential to understand the patterns of patient messages, what they write about, and what details they tend to include or leave out. Characterizing a large pool of messages into a structured set of categories reveals these patterns and could inform building automated tools that aim to reduce the communication burden. Thus, a structured representation is required to characterize patient messages. Such a characterization can help the design of artificial intelligence (AI) and natural language processing (NLP) tools tailored to portal communication, such as automated assistants that help patients draft more complete messages and help clinicians draft responses to reduce their workload, with the aim of making communication more efficient and reducing the back and forth communication it requires [4,5].

Prior work has described portal messages mainly by assigning *topic* labels. Early studies developed taxonomies of message content and applied them through manual or automated annotation [6–8]. More recent work has used large language models (LLMs) to label messages at scale. Steitz et al [9] classified 2.4 million messages into an expert-derived 11-category taxonomy and found that content concentrated in a few topics, with about half of messages addressing more than one topic, and Chang et al [10] used unsupervised LLM-based topic modeling with topic labeling to categorize messages. Topic labels show what a message is about, but not the details it contains, such as when a symptom began, which medication is involved, or what kind of appointment the patient wants. These details determine whether a clinician can act on a message or must first ask the patient for more information. Structured representations exist for other clinical text, including biomedical terminologies and concept extraction from clinical notes [11–13], but these were not designed for patient-authored messages, and existing portal-message schemas support topic analysis rather than this level of detail [14].

We treat this as an Event Extraction (EE) task [15,16], which is an NLP task that converts event-centric text into structured objects. Its subtask, Event Argument Extraction (EAE), extracts the details of each detected event [15,16]. Portal messages are largely event-centric, describing symptoms, medications, appointments, and similar occurrences. EE represents each occurrence as an event type, such as a symptom event, with a set of roles, which represents the details of the event, such as the symptom onset, all defined in advance in an ontology. Figure 1 illustrates this with a synthetic patient message annotated with its event types and roles. Because the ontology fixes the vocabulary, extracted details can be normalized, so that equivalent expressions such as “twice a day” and “BID” receive the same label and results from individual messages can be aggregated across a corpus. LLMs suit this task because they can extract clinical information from instructions alone, without labeled training data [17], and EE can be posed as a series of questions about each role, a formulation well matched to LLMs [18,19]. Our setting adds a practical constraint. Patient messages contain protected health information, so the framework must run on smaller open-source LLMs on a secure, virtual computing platform.

**Figure 1.**
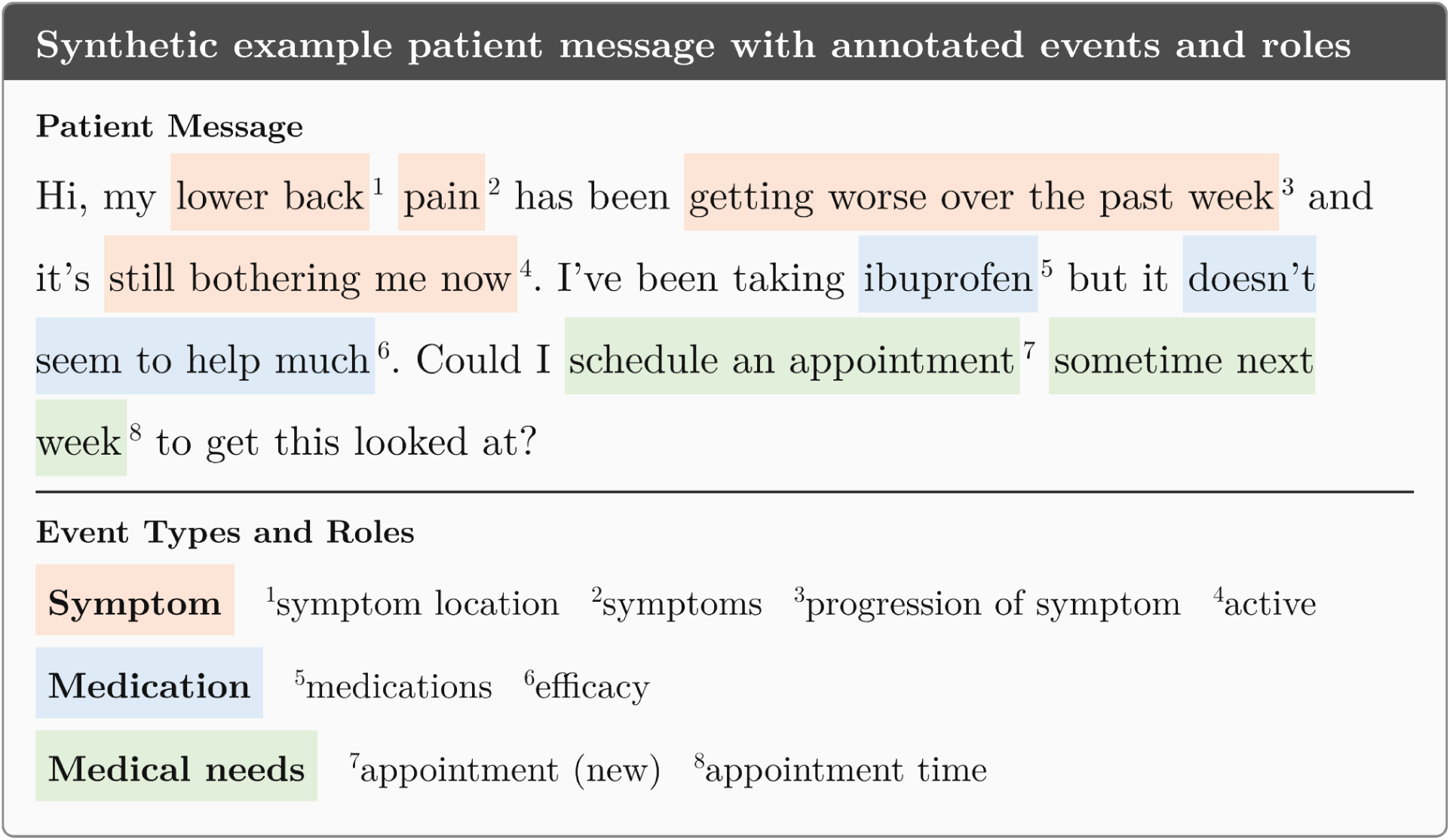
A synthetic patient message annotated with PortalEvent event types and roles. Highlighted spans mark the text that instantiates each role, color-coded by event type. The example illustrates three of the most frequently detected event types (symptom, medication, medical needs) and several of their common roles.

We introduce PortalEvent, an event ontology for patient portal messages that spans 8 event types and 71 roles. The ontology was built in three stages, (i) a literature review of existing healthcare communication schemas [8,20–22], (ii) LLM-assisted analysis of real patient messages to surface content the literature did not cover, and (iii) iterative review by NLP researchers and practicing clinicians. PortalEvent makes it possible to analyze patient portal messaging, for example, characterizing what patients write about, what clinicians include in their replies, and questions clinicians ask to obtain information a patient message lacks. In this work we focus on characterizing both patient messages and clinician follow-up questions.

We applied the framework to a sample of 10,000 real patient messages using three open-source LLMs, running locally on the secure computing environment, and the three models gave broadly consistent results. In the output of one of the three models, GPT-OSS-20B, medical needs (55.3%, 5,534/10,000) and symptom (52.7%, 5,267/10,000) were the most frequent events, and treatment response was the least frequent (10.9%, 1,091/10,000). Administrative and clinical content often appeared in the same message, with medical needs and symptom events appearing together in more than a quarter of messages. In a manual review by two independent annotators, at least 94% of detected events were correct for five of the eight event types, and the annotators agreed on 91% of 826 judgments about event types and 96% of 751 judgments about the extracted details (excluding treatment response, which was evaluated on a separate sample).

As an extrinsic evaluation, we leveraged the ontology to characterize clinicians’ follow-up communication on patient portals. When a message lacks information the clinician needs, the clinician often replies with a follow-up question, which delays resolution of the patient’s request and adds to clinicians’ portal-related workload. Across many messages, these questions show what information clinicians consistently need but patients tend not to specify in their message [4]. We therefore applied the PortalEvent ontology to characterize clinician follow-up questions in our dataset and surface common events and arguments that lead to followup questions. From clinician responses containing a question mark, we extracted 57,321 follow-up questions across 32,278 messages and labeled each with the event types and roles it asks about. Symptom was the most frequent target, accounting for 40.4% (23,163/57,321) of questions, followed by medication (20.2%, 11,560/57,321) and medical needs (20.2%, 11,551/57,321). Within each event type, questions concentrated on a few roles, most often the one that establishes the basic facts of the event, such as which symptom is present, which medication is involved, or whether a new appointment is needed. These patterns give an overall trend of what information clinicians seek from patients, which could inform tools such as automated follow-up question generation [4].

To summarize, this paper makes the following contributions:

- We introduced PortalEvent, an event ontology of 8 event types and 71 roles for patient portal communication, together with a training-free extraction framework deployable on open-source LLMs.
- We validated the ontology by applying the framework to 10,000 real patient portal messages, and characterizing the distribution of extracted event types and roles, showing that the ontology covers a broad range of content expressed through this channel.
- We applied the same ontology to clinician follow-up questions. From approximately 146,000 patient message-clinician response pairs, we identified 32,278 pairs in which the clinician response contained at least one follow-up question, which yielded 57,321 questions in total. We used the ontology to characterize these questions, and the distributional analysis provides a structured foundation for downstream applications, such as ontology-guided follow-up question generation that prompts patients to supply missing information before the clinician reads the message.

### Related Work

#### Event Extraction

Event extraction (EE) is an NLP task that converts event-centric text into structured objects. Classical formulations of EE decompose the task into two sub-problems: event detection (ED), which identifies which events occur in a passage, and event argument extraction (EAE), which identifies the details of each detected event. In contrast to open-domain information extraction [23,24], EE is grounded in a pre-defined event ontology [15,25]: domain experts specify a set of event structures that the extraction system is permitted to instantiate. Event types and roles are properties of the ontology, and the arguments that fill those roles are extracted from the document. As an example, Table 1 illustrates a symptom event, which is detected when a patient message describes a physical or mental symptom. The roles of this event capture details such as the symptoms reported, their location on the body, and how they have progressed over time. To illustrate the terminology of event extraction, the argument of the *progression of symptom* role for this symptom event is *worse*.

**Table 1.** Example event in the PortalEvent ontology: the *Symptom* event captures a patient’s report of a physical or mental symptom. Each role has an output space (BIN: binary, CAT: categorical, FREE: free text, MULTI: multiple values) and a set of arguments the role can take. The five roles shown are the most frequently sought in clinician follow-up questions. the full symptom event defines 15 roles.

| **Role** | **Type** | **Description** | **Example arguments** |
|---|---|---|---|
| symptoms | MULTI | The symptoms the patient reports. | “*lower back pain*”, “*headache*”, “*nausea*” |
| symptom_location | FREE | Where on the body the symptom occurs. | “*lower back*”, “*left knee*”, “*right temple*” |
| active | CAT | Whether the symptom is current, denied, or resolved. | active / negated / resolved |
| progression_of_symptom | CAT | How the symptom has changed over time. | better / worse / same |
| aggravator | FREE | Patients attributed factor that triggers or worsens the symptom. | “*after lifting boxes*”, “*since starting the new job*” |

EE has been specialized to different problem settings. Classical EE assumes that both events and their arguments are extracted directly from text spans [26,27]. This assumption simplifies extraction but limits the modeling of events whose arguments are implicit or scattered across non-adjacent passages. Recent work has used LLMs to relax this assumption: our prior work on DiscourseEE [16] extends EE to extract arguments that are implicit or distributed across the discourse. A recent survey argues that event extraction is still useful with LLMs, and event schemas and slot constraints serve as interfaces for grounding and verifying model output [28]. Schema-constrained extraction is also competitive with open models: fine-tuned mid-sized open-source LLMs outperform previously reported GPT baselines on document-level argument extraction under a strict schema [29]. Taken together, EE provides a framework for ontology-grounded characterization of events in a corpus, and LLM-based methods extend its reach to domains in which arguments do not align with simple text spans.

#### Characterization of Patient Portal Messages

Patient message corpora are an access-restricted data source, and existing work on characterizing such corpora has relied primarily on taxonomy-based annotation. Cronin et al [6] developed a taxonomy of consumer health information needs, annotated a corpus of 1,000 patient-generated portal messages, and evaluated rule-based and machine-learning classifiers for detecting taxonomy categories. Shimada et al [7] coded the content of secure message threads in Veterans Affairs primary care, reporting how often patients requested medication refills, scheduling, help with health issues, or referrals. Heisey-Grove et al [8] developed a theory-based taxonomy for patient- and clinician-generated secure messages and applied it through manual annotation.

More recent work has applied automated NLP methods to message characterization. Mastorakos et al [30] used a named-entity recognizer aligned to Unified Medical Language System (UMLS) concepts to characterize approximately 3,000 messages. Sulieman et al [31] analyzed the medical terms, topics, and linguistic features of patient portal messages and their association with hospital readmission. Chang et al [10] used unsupervised LLM-based topic modeling to identify common topics in cancer center patient messages. Steitz et al [9] classified 2.4 million medical advice messages with a zero-shot LLM pipeline and an expert-derived 11-category taxonomy, finding that content concentrated in a few topics and that about half of messages addressed more than one topic. A related line of work extracts clinical content from clinical text rather than assigning topic labels: Steinkamp et al [32] defined a symptom-extraction task with an annotated corpus and supervised models for extracting symptoms from unstructured clinical notes.

Two limitations of this prior work motivate our study. First, existing taxonomies are underspecified for structured analysis: they categorize messages at the topic level but do not provide an event-centric representation of message content. The ontology introduced in this work specifies 71 roles across 8 events, supporting extraction at a finer granularity than topic categorization. Second, most prior work relies on training data drawn from a single institution’s private corpus, which constrains generalizability. We adopt a training-free LLM-based extraction approach that can be applied to a private corpus without site-specific training. Whether the resulting distributions transfer across sites is an open question that our single-site evaluation cannot fully address.

#### LLM-Based Ontology Construction

Beyond message characterization, our methodology also draws on a growing body of work that uses large language models to reduce the manual cost of building and populating ontologies. The LLMs4OL paradigm [33] framed ontology learning as a set of LLM-addressable subtasks (term typing, taxonomy induction, and non-taxonomic relation extraction) and evaluated zero-shot prompting across general-purpose resources such as WordNet, GeoNames, and UMLS. Subsequent studies examined how closely LLM-produced ontologies match human modeling: Saeedizade and Blomqvist [34] reported that GPT-4-class models approach the quality of novice human ontology engineers on generation tasks. A recent systematic review of 36 papers finds that LLMs in ontology engineering mainly act as ontology engineers, domain experts, and evaluators, and that most applications are in the life sciences and healthcare [35].

A second group of methods targets ontology population, in which an existing schema is held fixed and the LLM extracts schema-conformant instances from text. Norouzi et al [36] populated a modular domain ontology with an LLM guided by the schema in its prompt, recovering up to roughly 90% of ground-truth triples, with performance remaining similar across prompting and retrieval strategies when the schema was supplied. Pipeline systems such as OntoKGen [37] and the framework of Kommineni et al [38] combine LLM prompting with iterative human or chain-of-thought guidance to extract an ontology and then populate a knowledge graph, and benchmarks such as Text2KGBench [39] evaluate whether LLM-generated facts conform to a target schema.

Although healthcare is among the most common application domains for this work [35], its targets are typically formal domain knowledge, such as concepts, taxonomies, and relations, rather than the content of patient communication. We adapt its two central ideas to clinical patient-message characterization: we use LLM-assisted induction to construct candidate event types and roles from a message corpus (the Ontology Construction section), which domain experts then refine into a fixed ontology, and we then apply the same schema-guided, training-free strategy to characterize message content at the event and role level.

#### LLM-Based Ontology Extraction from Clinical Text

Closely related to ontology construction is the use of LLMs to *extract* schema-conformant information from clinical text, the task our framework performs. Agrawal et al [17] showed that general-purpose LLMs perform well at zero- and few-shot information extraction from clinical notes without domain-specific training, establishing training-free prompting as a viable alternative to supervised clinical extraction pipelines. Builtjes et al [40] found that locally run open-source LLMs with schema-validated structured output matched or outperformed a fine-tuned baseline on 17 of 28 clinical extraction tasks, with models around 14B parameters performing well overall. Subsequent work has paired this capability with fixed domain ontologies to produce structured clinical knowledge. Cao et al [41] introduced AutoRD, an end-to-end system that extracts rare-disease entities and relations from medical text using ontology-enhanced LLMs, reporting gains in relation extraction over the baseline LLM. Alhassan et al [42] combined generative models with sentence transformers to recognize and normalize Human Phenotype Ontology mentions, finding that exact span extraction was not always necessary for successful normalization to ontology concepts.

Our extraction task is structured as event-argument extraction, in which each detected event is associated with a set of typed roles. This connects our framework to recent work on generative event-argument extraction [16,43,44], which we adapt to short, patient-authored portal messages. Unlike the disease-knowledge systems above, which target formal entities and relations drawn from medical literature or clinical reports, our work extracts communicative events and their roles from messages written by patients. It shares their reliance on a fixed, expert-defined schema and training-free LLM extraction, but applies the paradigm to a different setting and a different unit of analysis.

## Methods

### Ontology Construction

In this section we describe the methods used to construct the PortalEvent event ontology schema, which is the set of event types, roles, and definitions that characterize information appearing in patient portal messages. Figure 2 gives an overview of the construction stages. Each stage is explained. The PortalEvent ontology schema was developed in three stages, namely *(1)* a literature review of existing healthcare communication taxonomies, *(2)* an automatic ontology-induction step over a corpus of real patient messages, and *(3)* iterative expert review by NLP researchers and three clinicians.

**Figure 2.**
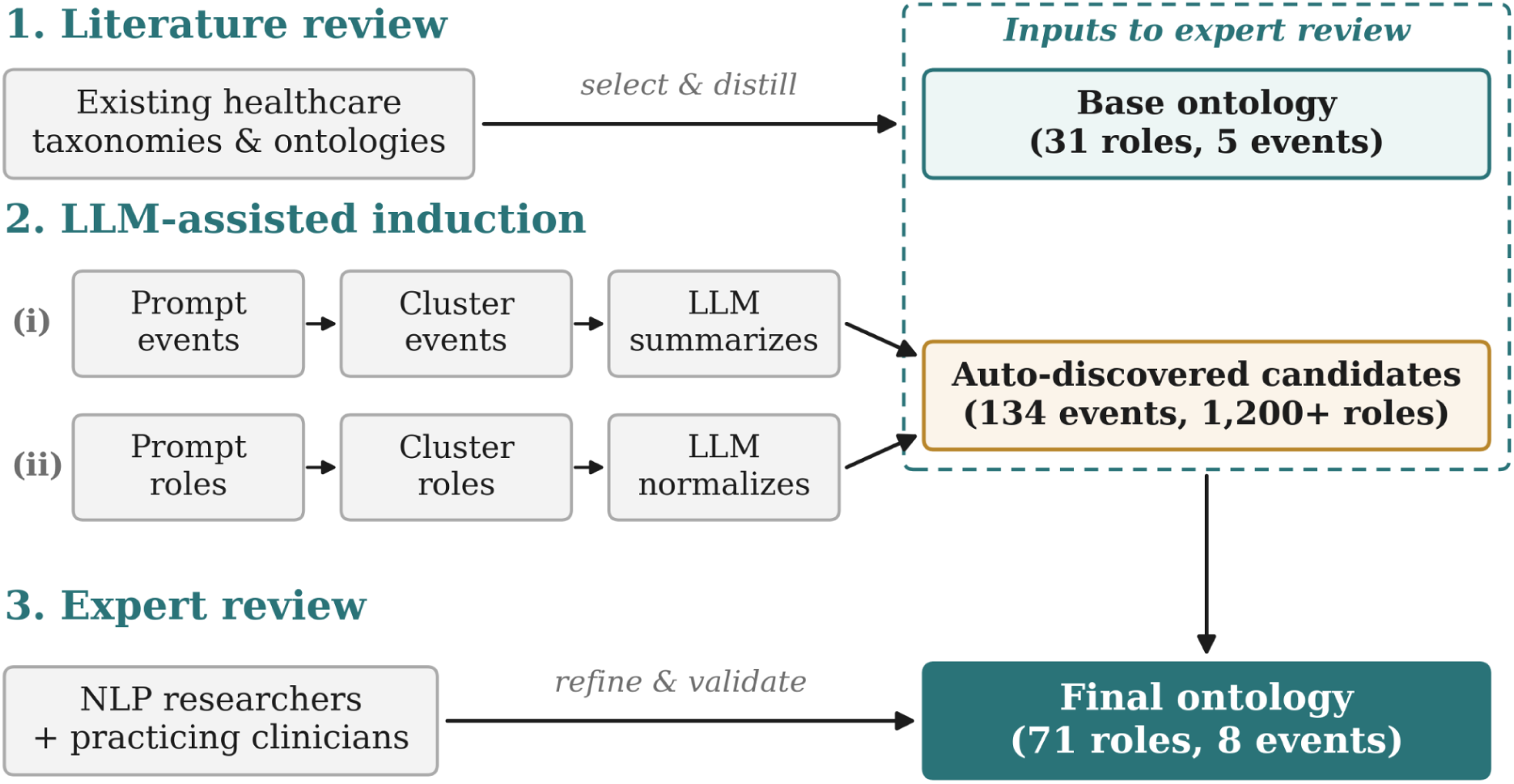
Workflow Pipeline of Ontology Construction.

#### Literature Review

During the first construction stage, we reviewed existing taxonomies and schemas of healthcare communication available at the time of construction [6–8,20–22]. From these sources, we selected the four that fit our event centric scope [8,20–22]. We then combined them into a unified ontology by reducing redundancies, merging concepts, and restructuring the terminology into an event-centric framing, in which each category became an event type with a set of roles describing its details. This stage proposed 31 roles across 5 event types.

#### LLM-Assisted Induction

To cover content missing from the literature-derived ontology, we applied an LLM ontology-induction approach following prior work on LLM-based schema induction [45,46]. Given a corpus of 1,000 real patient messages, we prompted an LLM to propose event types and roles from each message. An illustrative output of some LLM proposed events is shown below (Figure 3).

**Figure 3.**
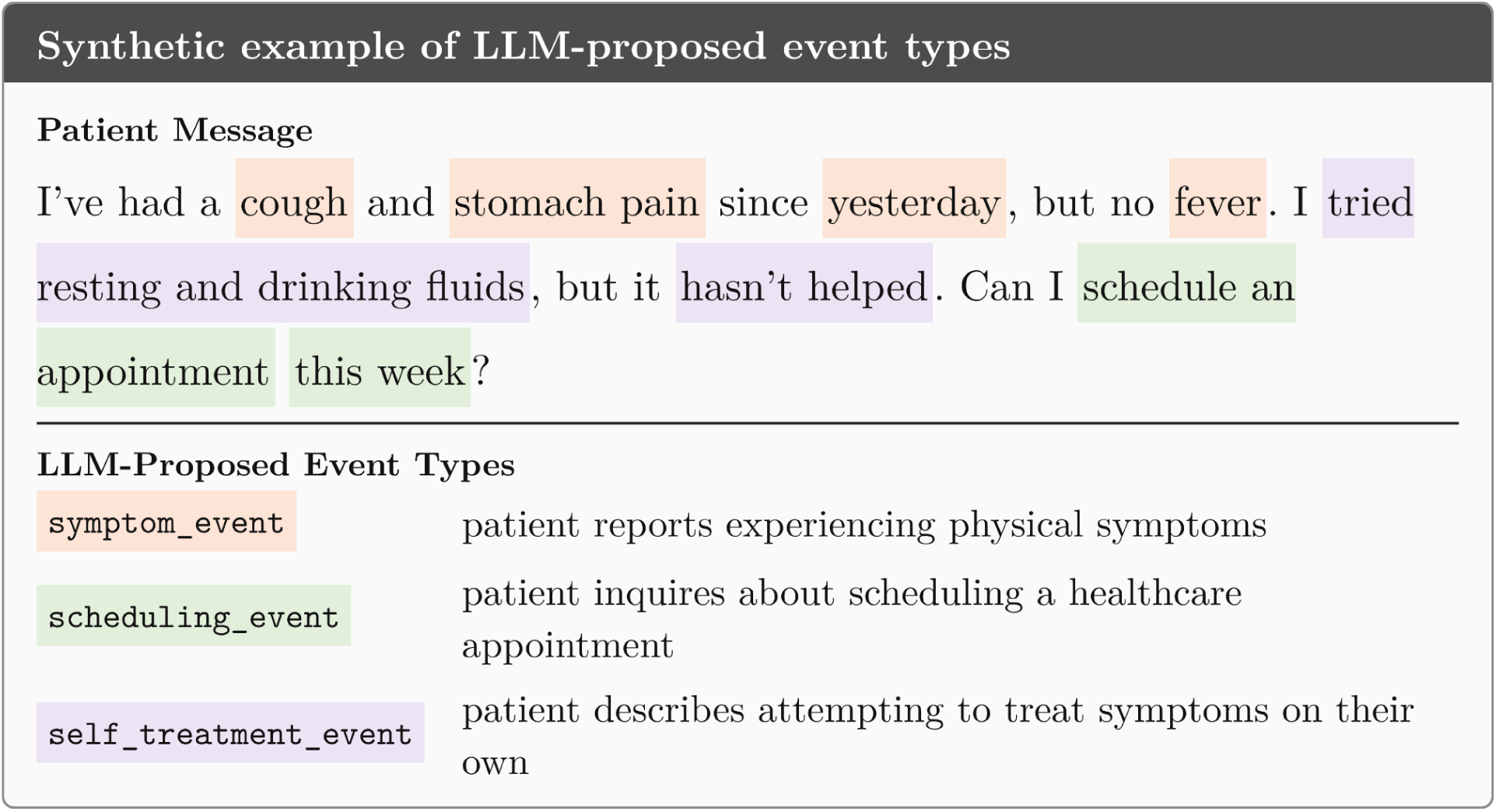
Synthetic example of LLM-proposed event types.

The LLM proposed events from the corpus were grouped through hierarchical clustering [47] over sentence-transformer embeddings of the event names and descriptions [48]. Each cluster was then summarized by an LLM into a canonical event title that represents the cluster’s members. For example, a cluster containing *scheduling_event*, *appointment_event*, and *reschedule_event* was summarized under a single canonical event type called *medical_needs*. Then, a similar procedure was applied to propose roles within each canonical event type. For every message detected as containing an proposed event type, the LLM was prompted to output the set of proposed roles, their arguments, and a brief description for each role. An illustrative output is shown below (Figure 4).

**Figure 4.**
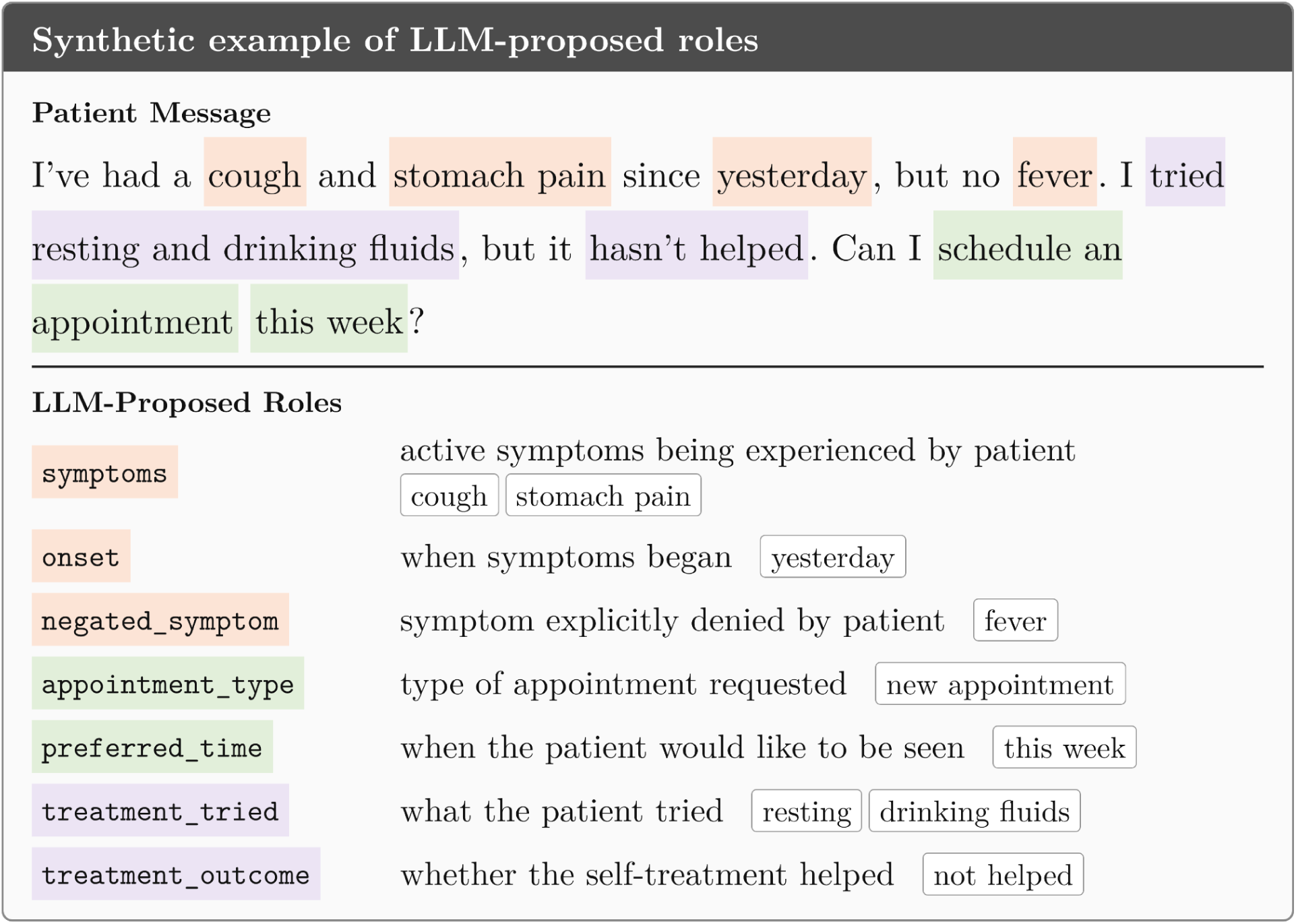
Synthetic example of LLM-proposed event roles.

Proposed role names and descriptions were clustered and summarized using the same procedure, which produced a candidate set of roles per event type. Applied to a corpus of 1,000 real messages, this process yielded 134 candidate events and over 1,200 candidate roles. The raw output was not adopted directly for two reasons. First, many proposed event types were too narrow to serve as ontology entries. For example, distinct entries for blood pressure events or cancellation events fit naturally under more abstract categories such as diagnostic test or scheduling. Second, scaling this process to larger corpora has limits. When a cluster exceeds the LLM context window, the normalization step requires sub-sampling cluster members, which can result in information loss. The induced ontology was therefore treated as an additional signal, alongside the literature-review baseline. A doctoral researcher working in medical NLP and fluent in portal events and workflow reviewed the induced ontology to discover new events and roles. This review led the team to add events and roles for self-reported emergency department and urgent care visits, and for arguments related to mental health, that were not surfaced by the literature review.

#### Clinician review of the ontology

The induced ontology was validated by three clinicians (MDs). The first, TEB, a family physician (MD MBA MSc) at Dartmouth Hitchcock Medical Center (DHMC) with over 20 years of clinical practice, who is also Associate Chief Research Officer for Informatics and Vice Chair for Research in the Department of Community and Family Medicine, reviewed and revised the ontology schema during construction through five iterations. Then he conducted the ontology coverage annotation. The second, SKL, an associate professor of Medicine, Health Policy and Clinical Practice at Dartmouth Geisel School of Medicine with over 25 years of experience, reviewed the ontology schema and provided feedback on the coverage and definition of events and roles. The third, MK (MD, PhD), a family medicine physician at DHMC, helped with reviewing the ontology schema and provided feedback on the coverage and definition of events and roles. The clinicians identified gaps in the ontology, and they determined which abstract categories were important for representing portal-message content, and revised role definitions over a few iterations. We finalized the ontology schema based on their feedback.

After the schema was finalized, we ran a coverage check to test whether the events could express what patients actually write. A random sample of 30 messages was drawn from the corpus and reviewed by TEB, who saw each message alongside the complete list of event types with their definitions and sample roles. For each message, the clinician marked the message could be expressed by an existing event type, or otherwise proposed the missing type in free text. The other clinicians reviewed the pre-defined ontology schema. No new event types or roles were proposed. The feedback instead concerned event role definition boundaries. Eight definitions in either role name or role description were clarified or modified(i.e., organ system, symptom type, symptom severity, prescription request, financial concern, personnel referrals, complaints, response).

The final ontology spans 8 event types and 71 roles. The event types and their definitions are listed in Table S1 in Multimedia Appendix 1, and the full set of roles is listed in Table S2 in Multimedia Appendix 1.

#### Role Output Space Design

The output space for each role represents the set of arguments it can take. Existing event extraction systems define arguments either as text spans [15,25] or generatively [16]. Both yield unstructured free text values that are difficult to aggregate across a corpus when the underlying information is heterogeneous. During the role extraction stage, if role argument were defined as text spans or generated freely, text spans from patient messages such as “twice a day,” “BID,” and “every 12 hours” would be extracted as three distinct values rather than one, so it would be challenging to aggregate the results reliably. Therefore, we defined normalized output space for applicable roles. We refer to a role’s output as *normalized* when the set of arguments are mapped to a closed, predefined set rather than copied verbatim from the message. Normalization is what allows corpus-level aggregation. In PortalEvent, this was implemented through the argument type of each role. Each role was assigned one of four output space types. Binary and categorical roles have a closed set of possible arguments, their outputs are fully normalized. The medical needs event, for example, contains eleven roles, ten of which have a binary or categorical output space. Rather than collapsing appointment-related content into a single role with a free-text value describing the nature of the request, we defined four separate binary roles for new, rescheduled, canceled, and confirmed appointments. The resulting outputs can be aggregated directly at the corpus level, which lowers the cost of producing macro-level statistics. Some roles cannot be normalized. The factors that aggravate a symptom, for example, are open-ended and cannot be reduced to a closed label set. For these roles we retained a free-text output, since a clinical collaborator identified them as substantively important and they remain usable for modeling at smaller scales. Roles whose arguments cannot be reduced to a closed set, such as the list of symptoms a patient reports, are defined as free text or multi-value free text.

### Event Detection and Role Argument Extraction

#### Data and Experimental Setup

We applied the final PortalEvent ontology to a corpus of patient portal messages and paired clinician responses from Dartmouth Health, an academic health system in New Hampshire, United States, introduced in prior work [4,5]. The messages were sourced from the hospital’s primary care service through its electronic health record (EHR) portal messaging platform, an asynchronous channel in which patients and clinicians discuss a wide variety of health issues including symptoms, medication efficacy, treatment planning, and scheduling logistics. The corpus comprises approximately 146,000 patient-initiated messages, each paired with a written response from a clinician. We applied the LLM extraction framework to a random sample of 10,000 patient messages from this corpus and report aggregate statistics over the extracted events and role arguments. Table 2 summarizes the characteristics of this sample.

**Table 2.**
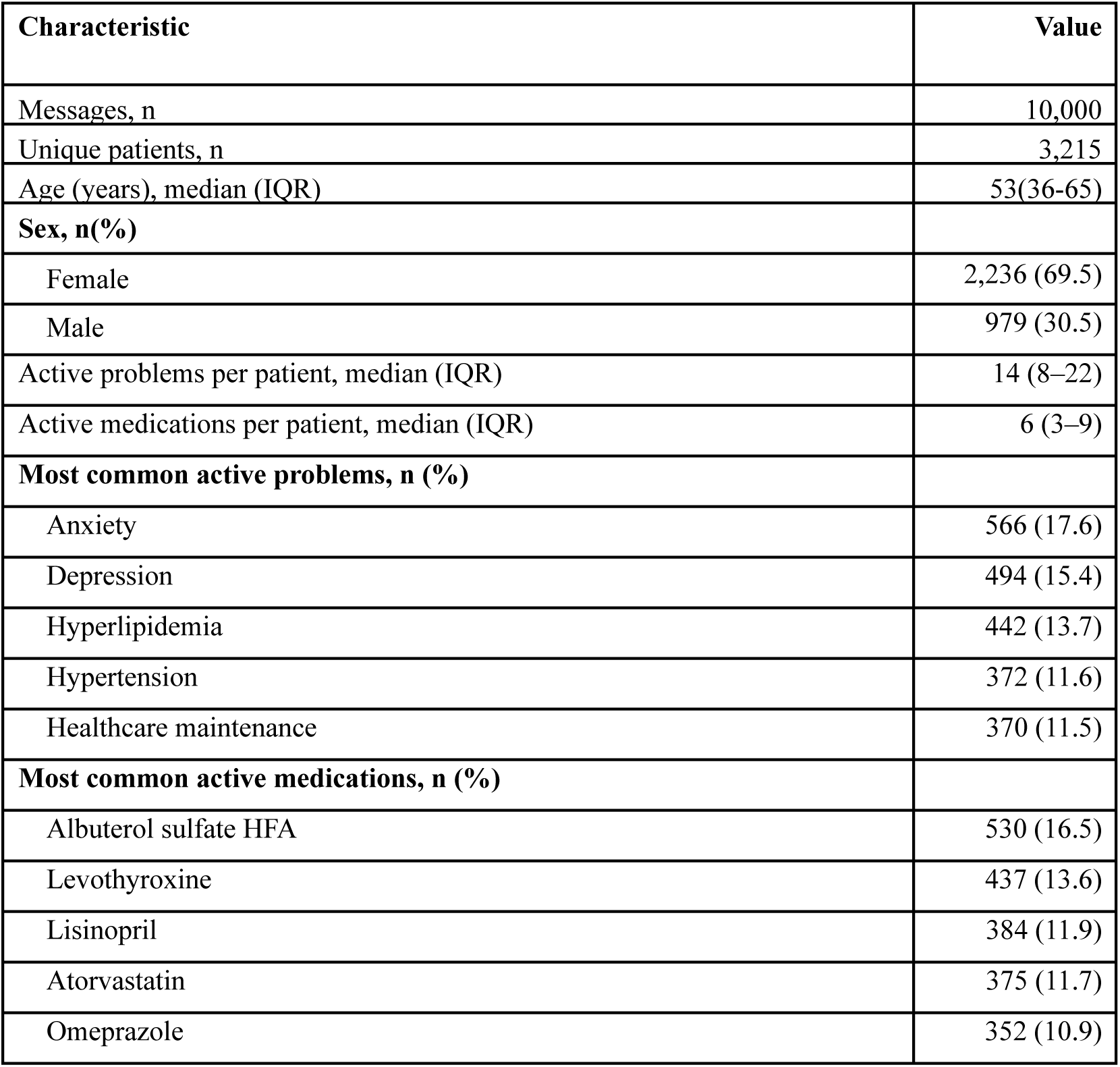
Characteristics of the 10,000-message sample (January 2020–December 2024). Percentages for sex, problems, and medications are of the 3,215 unique patients. Age was recorded in ranges and calculated from range midpoints. IQR: interquartile range.

The corpus is hosted by Dartmouth College on tiCrypt (Tera Insights LLC, Gainesville, FL) virtual compute environment with NIST SP 800-171r3 and CMMC 2.0 Level 2 security compliance. The system has no external network access. All processing of real patient messages ran on a secure, offline computing environment. The platform provides NVIDIA A40 graphics processing unit (GPU) with 48 GB of video memory, which restricts the framework to 4-bit quantized language models of at most 32B parameters. We ran the framework with three open-source models spanning general-purpose and medical-domain families, GPT-OSS-20B, MedGemma-27B, and Qwen3-32B, to assess whether the extracted structure depended on the choice of the LLM models. All three ran locally within the secure environment, consistent with the privacy constraints of the corpus. Unless otherwise noted, we report detailed statistics using GPT-OSS-20B and use the remaining two models to assess consistency across LLM extractors. The framework is training-free.

#### Applying PortalEvent Ontology to Portal Messages

The extraction process consists of two steps: event detection followed by role extraction.

*Event detection.* In PortalEvent, the aim of event detection is to extract event type labels from patient messages. Given a patient message, we prompted an LLM model to extract the event types that are present in the message. For example, a patient message could contain a symptom event, medication event, and medical needs event. We formulated this as a multi-class multi-label classification problem. To do this, we distinguished two detection patterns based on the relationship between an event type and its roles.

*Bottom-up event types*: patient context, medical needs, logistics, and social. The presence of these event types cannot be inferred directly from the event type name, because each covers many different kinds of content. For example, logistics includes record requests, insurance questions, and IT problems, so a single question such as “Does the message contain a logistics event?” is too broad to answer reliably. Instead, we first asked the LLM about each role directly, for example, “Does the patient request a copy of their medical record?” If any role was present, then its corresponding event type was present.

*Top-down event types*: symptom, medication, diagnostic test, and treatment response events. In contrast, the presence of these event types can be inferred directly from the event type name. We first asked the LLM whether the event type was present, for example, “Does the message describe a symptom?” When the answer to these questions was yes, we then extracted the corresponding roles, such as the symptom’s severity or onset for a symptom event.

Figure 5 illustrates the two patterns.

**Figure 5.**
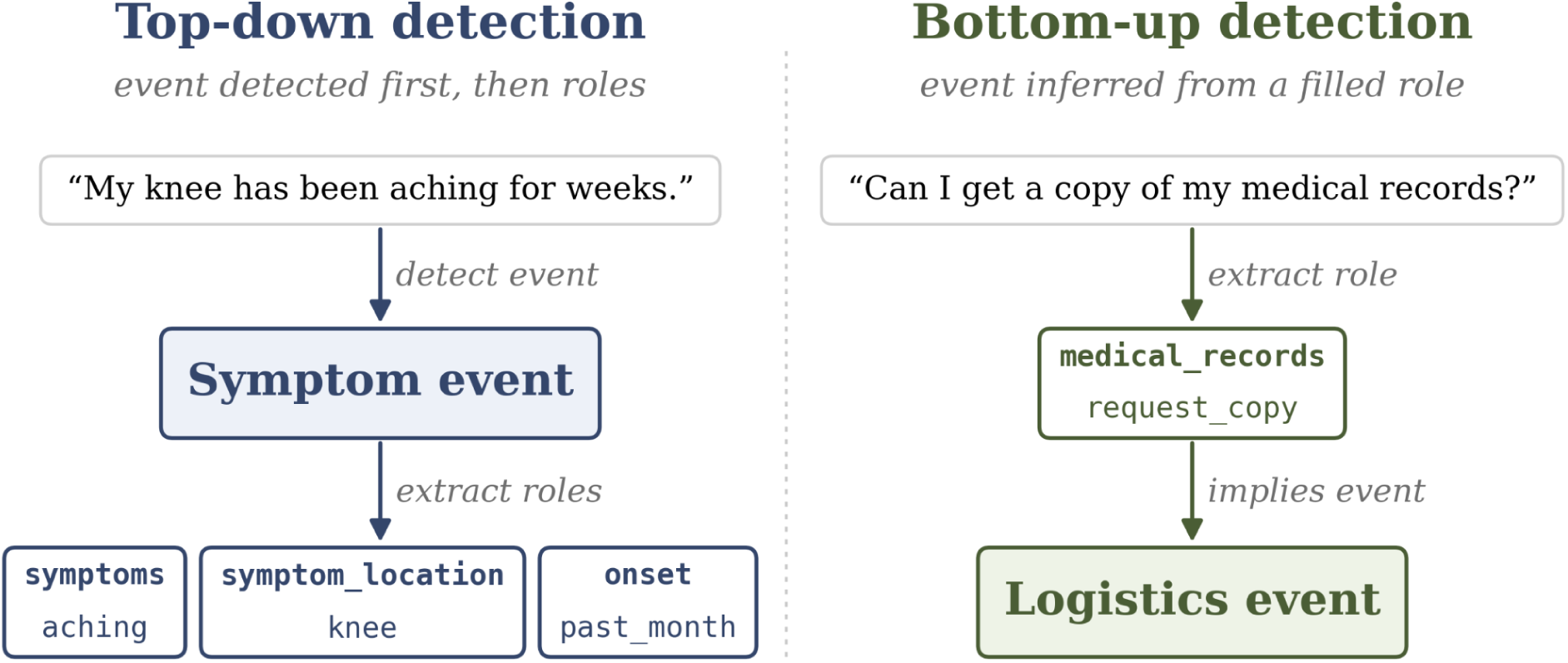
Two event detection patterns in the ontology. Top-down events are detected before role extraction, and bottom-up events are inferred whenever any of their roles is present.

*Role argument extraction:* For top-down event types, we extracted the arguments of each role after the event type was detected. For bottom-up event types, the role prompts used during event detection provided the arguments. To extract role arguments without training and under constrained GPU resources, we adopted a question-answering formulation of event argument extraction (EAE) [18,19]. Rather than prompting the LLM with the full ontology and asking it to extract all role arguments under a detected event type, we used a separate prompt per role (e.g., “is the symptom getting better, worse, or staying the same?”).

#### Prompt Design

Extraction of each role argument used a dedicated prompt with a fixed instruction, a small set of in-context examples, and a constrained output schema. An example prompt for the *active* role of the symptom event is shown below (Textbox 1).

Textbox 1. Argument Extraction Prompt for “Active” in the “Symptom” event.

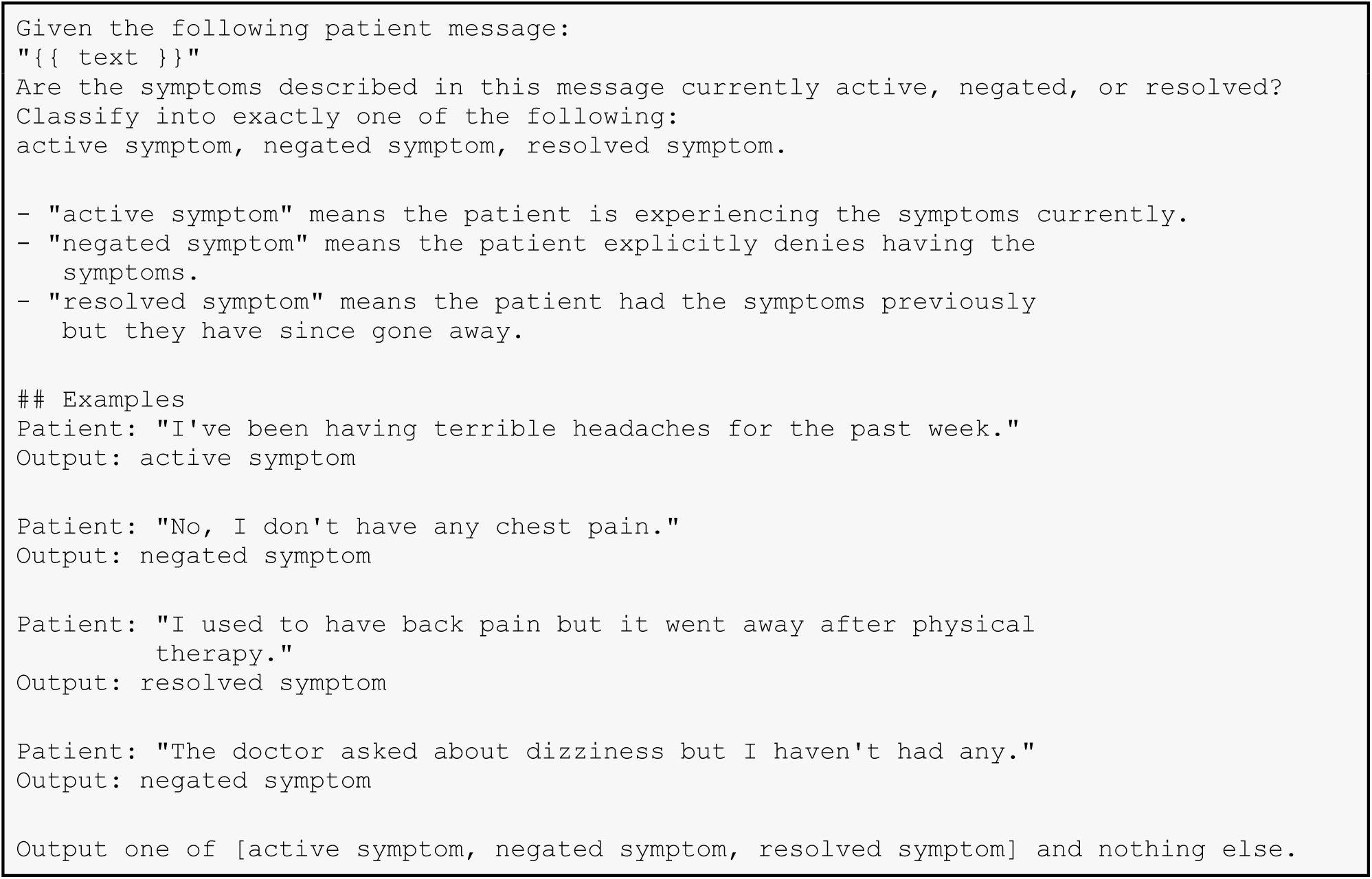

The prompts contain no protected health information, and all example messages shown in this paper are synthetic.

#### Expert review of LLM Extraction Output

To evaluate the quality of the characterizations of patient messages, which are event types and role arguments extracted by the LLM from patient messages, two doctoral researchers working in clinical NLP and familiar with patient portal messaging and clinical workflows reviewed the extractions produced by GPT-OSS-20B for precision, the proportion of extracted event types and role arguments that were correctly assigned by the LLM. Both annotators independently reviewed the same items, drawn from a random sample of 30 messages, and annotated (i) whether each detected event type was correct (detection precision) and (ii) among correctly typed items, whether the extracted argument was correct (argument accuracy). We focused on precision evaluation because the result statistics reported in this paper are built from the events and role arguments the LLM models extracted, so the precision of those extractions indicates the quality of the reported counts. Precision is also a more efficient way to measure, since manual annotation of recall of the extractions for patient messages is substantially more costly because it may require annotators to independently annotate every event type and role to determine which event types or role arguments were missing.

#### Ontology-guided Characterization of Clinician Follow-Up Questions

When a patient message lacks information the clinician needs, the clinician often seeks additional information through follow-up questions. Across a corpus, these questions indicate what information clinicians consistently need but patient messages tend to omit. We applied the PortalEvent event ontology to characterize clinician follow-up questions. We constructed a follow-up question subset, prompted an LLM model to label each question with the event types and roles it asks about, and aggregated the labels across the corpus. A synthetic example of clinician follow up question characterization is shown below. (Figure 6)

**Figure 6.**
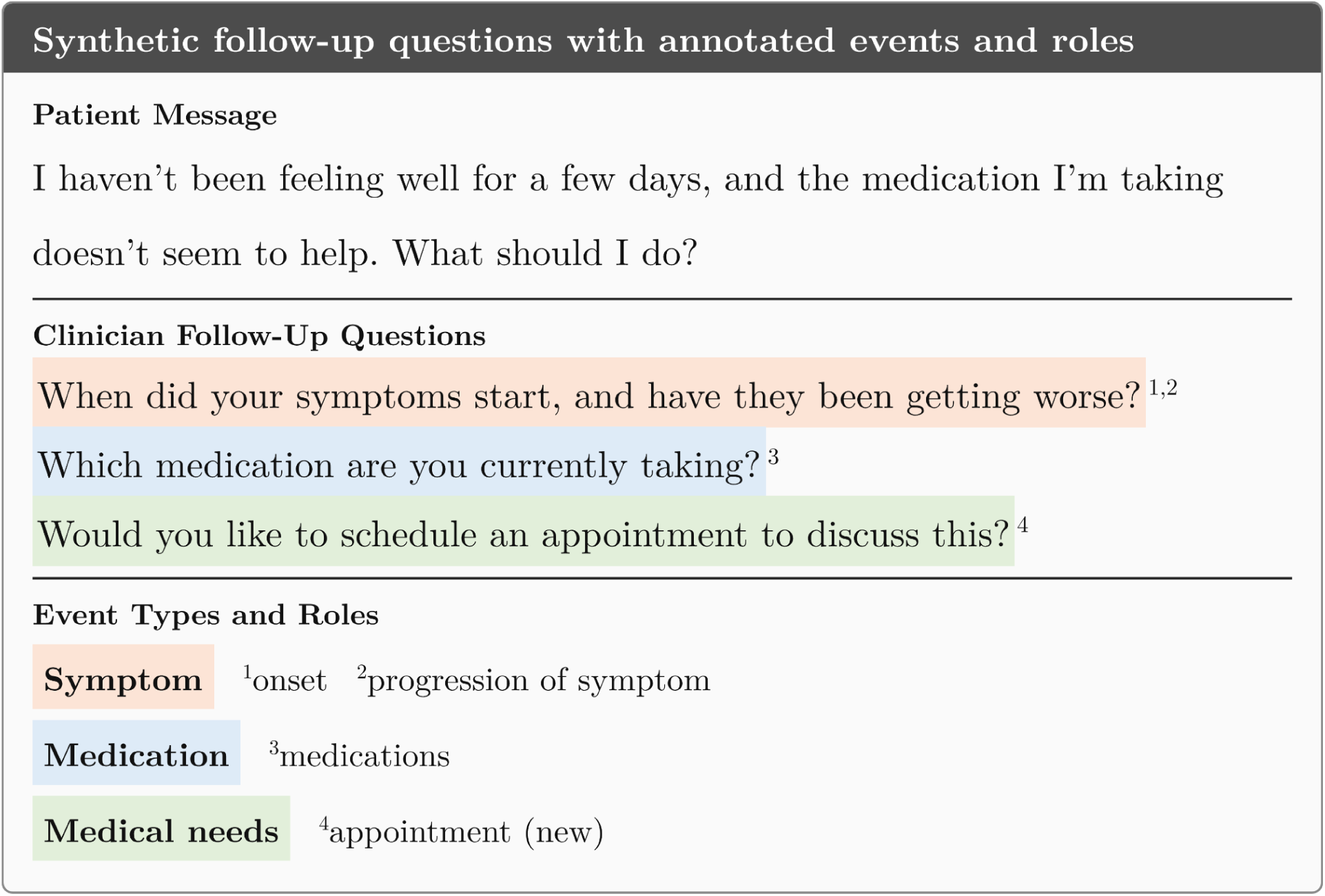
Synthetic clinician follow-up questions labeled with the event types and roles they ask about.

##### Follow-up question subset construction

We constructed the subset from the corpus containing approximately 146,000 patient messages. We first identified candidate messages, those whose clinician response contains at least one sentence ending in a question mark, which yielded 33,625 messages. From each candidate response, we then used Qwen3-8B to extract the individual follow-up questions. The model extracted at least one follow-up question from 32,278 of these messages. In the remaining samples with a question mark but that did not have any follow-up questions extracted, the question mark typically belonged to a tag or rhetorical question rather than a clinical follow-up question. The resulting subset, summarized in Table 3, contains 57,321 follow-up questions across 32,278 messages, with a mean of 1.8 questions per message.

**Table 3.**
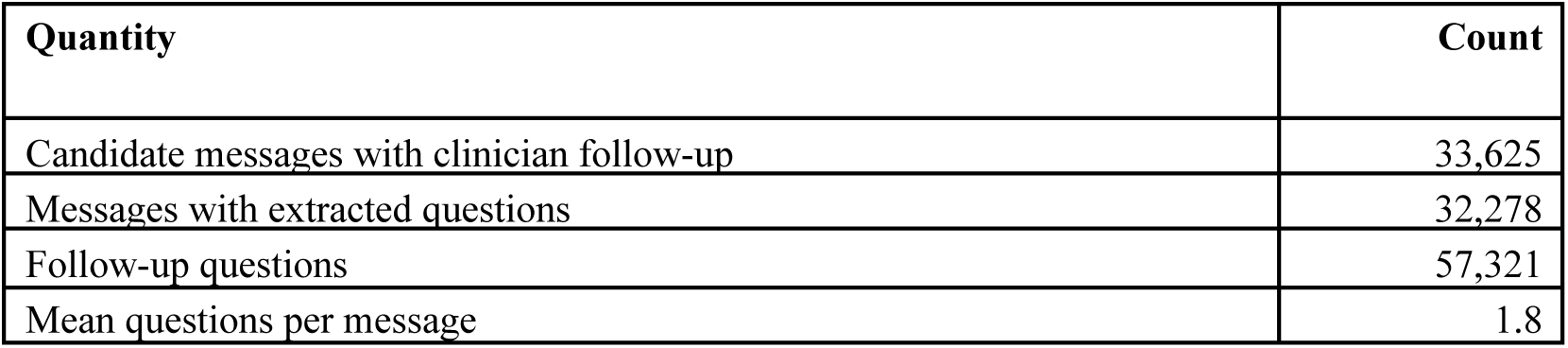
Follow-up question subset. *Candidate messages* are those whose clinician response contains at least one sentence ending in a question mark. A large language model then extracted individual follow-up questions, 32,278 messages with at least one. The mean is computed over those 32,278 messages.

##### Two-stage classification

We prompted an LLM model to characterize clinician follow-up questions in two stages. In the first stage, the model took in the follow-up question and the patient message as context, and the full list of eight event types, and the model output every event type the question asks about. In the second stage, for each selected event type, the model saw all roles within that type and selected the role the question asks about. Because a single question may ask about more than one event type or role, labeling was multi-label at both stages. A synthetic example showing the classification process is shown below. (Figure 7)

**Figure 7.**
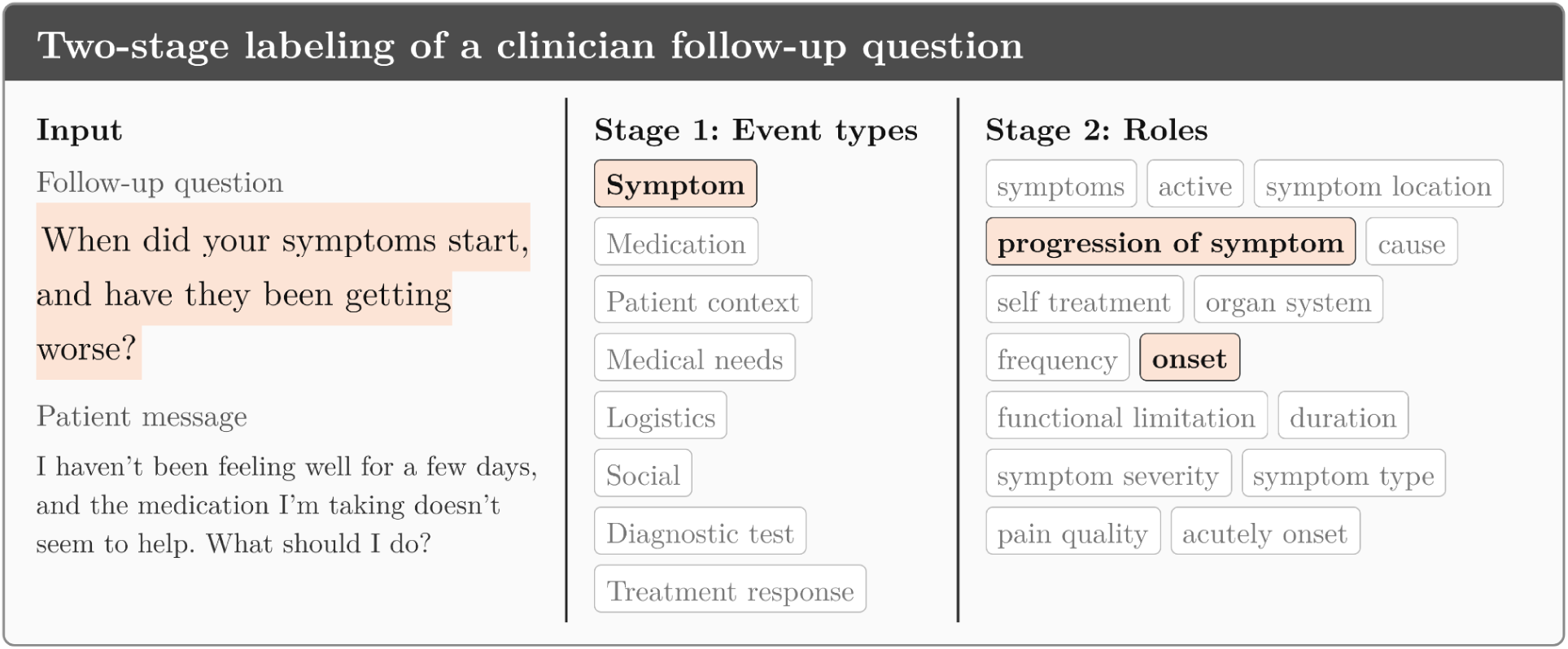
Two-stage labeling of a synthetic clinician follow-up question. The model first selects the event types the question asks about, then the roles within each selected event type.

##### Aggregation

Aggregating the labels across the corpus produced two distributions, the share of questions targeting each event type (the Event Types Targeted by Clinician Follow-Up Questions section), and, within each event type, the share of questions targeting each role (the Roles Targeted by Clinician Follow-Up Questions section).

##### Expert review of clinician followup questions characterization

A doctoral researcher working in medical NLP evaluated the characterization of clinician follow-up questions produced by Qwen3-32B, on a random sample of 200 questions. For each question, the annotator judged whether each assigned event type was correct and, among the correctly selected event type, whether the assigned role was also correct. Judgments covered all eight event types but were unevenly distributed due to random selection of samples. This evaluation measured precision. As with patient messages, the distributions of event types and roles reported for clinician follow-up questions are built from the labels the model assigned, so precision indicates how reliable these distributions are.

### Ethical Considerations

The Dartmouth Health Institutional Review Board approved this study (STUDY00033227). The patient portal message corpus contains sensitive patient information that cannot be publicly shared. All processing was carried out exclusively within a secure computing environment with no external network access, and the data were accessed only by personnel listed on the approved protocol, all of whom had completed Health Insurance Portability and Accountability Act (HIPAA) training. Clinicians who reviewed only the ontology schema did not access patient data and were outside the scope of the protocol.

The study itself poses no direct risk to human subjects. However, operational deployment of ontology-based extraction in real clinical workflows requires care. Errors in extracted event types or roles could misrepresent the content of a patient message, with potential downstream effects on triage, response drafting, or any system that relies on the extracted output. We therefore recommend that any such deployment include human review of extraction outputs, validation on the patient population at the deployment site, and ongoing monitoring of extraction quality over time.

## Results

### Event Detection and Role Extraction Results

Table 4 reports the distribution of messages in which each event type was detected, across all three extraction models. The trends were broadly consistent across models: the relative ordering of event types was mostly preserved, indicating that the ontology’s event structure is likely recoverable independently of the specific extractor model. One notable divergence was the social event, which Qwen3-32B detected less often (31.9%) than GPT-OSS-20B (40.4%) or MedGemma-27B (40.3%).

**Table 4.**
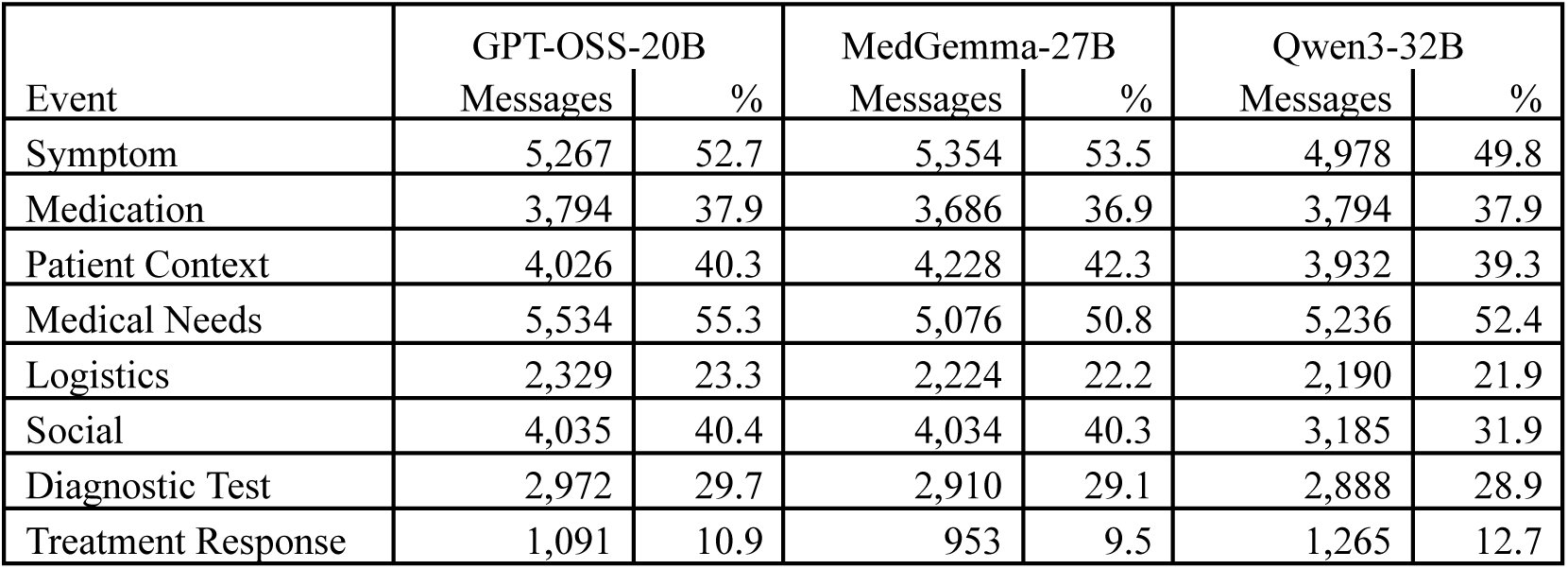
Event detection rates on the 10,000-message sample across three extraction models: GPT-OSS-20B (general-purpose), MedGemma-27B (medical-domain), and Qwen3-32B (general-purpose). For each event type and model, the table shows the number of messages in which the event was detected and the corresponding percentage of the sample. The denominator is 10,000 messages.

|  | GPT-OSS-20B |  | MedGemma-27B |  | Qwen3-32B |  |
|---|---|---|---|---|---|---|
| Event | Messages | % | Messages | % | Messages | % |
| Symptom | 5,267 | 52.7 | 5,354 | 53.5 | 4,978 | 49.8 |
| Medication | 3,794 | 37.9 | 3,686 | 36.9 | 3,794 | 37.9 |
| Patient Context | 4,026 | 40.3 | 4,228 | 42.3 | 3,932 | 39.3 |
| Medical Needs | 5,534 | 55.3 | 5,076 | 50.8 | 5,236 | 52.4 |
| Logistics | 2,329 | 23.3 | 2,224 | 22.2 | 2,190 | 21.9 |
| Social | 4,035 | 40.4 | 4,034 | 40.3 | 3,185 | 31.9 |
| Diagnostic Test | 2,972 | 29.7 | 2,910 | 29.1 | 2,888 | 28.9 |
| Treatment Response | 1,091 | 10.9 | 953 | 9.5 | 1,265 | 12.7 |

The LLM models were not able to detect any event types in some messages. For example, 253/10,000 for GPT-OSS-20B, 446/10,000 for MedGemma-27B, 387/10,000 for Qwen3-32B. We manually reviewed some random samples and found that many were short messages, and some messages contained at least one valid event type that the LLM model failed to detect.

We report the remaining statistics using GPT-OSS-20B. Medical needs was the most frequent event, detected in 55.3% of messages, followed closely by symptoms at 52.7%. Treatment response was the least frequent, detected in only 10.9% of messages. The remaining five event types were between 23.3% (logistics) and 40.4% (social).

Figure 8 shows that some pairs of event types co-occur in the same message. The strongest co-occurrence of two different events was between medical needs and social events (27.3%), followed by medical needs and symptom (26.2%), patient context and symptom (24.9%), patient context and medication (24.8%), and symptom and medication (24.1%). These pairs show that clinical and administrative content often appear in the same message, as in the example in Figure 1, where a symptom report is accompanied by a medication question and an appointment request. Figure S1 in Multimedia Appendix 2 presents these co-occurrences as a flow diagram.

**Figure 8.**
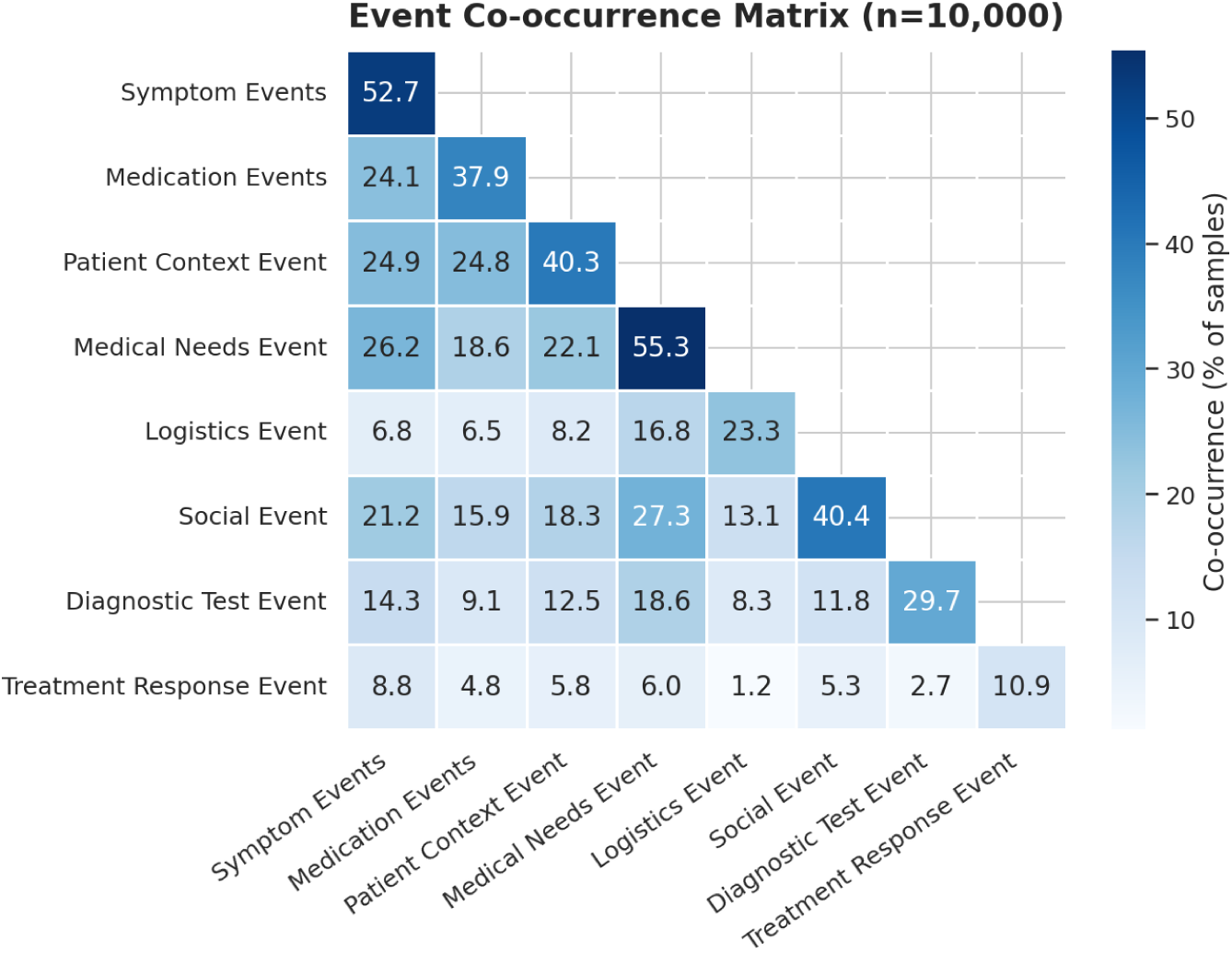
Co-occurrence matrix of the eight event types across the 10,000-message sample. Diagonal cells represent the marginal detection rate (percentage of messages in which each event type was detected), off-diagonal cells give the percentage of messages in which both events were detected together. All values are expressed as a percentage of the sample. For example, 24.1% of messages contain both symptom and medication events and 24.8% of messages contain both medication and patient context events.

Logistics showed a different pattern. Its co-occurrence with symptom, medication, and patient context fell below 10%. Treatment response, the rarest event overall, had the lowest co-occurrence across the matrix, with all of its off-diagonal cells below 9%.

Table S3 in Multimedia Appendix 2 reports the corpus-level characterization produced by the framework. The table includes binary, categorical, and multi-value roles, whose arguments are normalized and can be counted. Free-text arguments may vary in wording, so summarizing them across 10,000 samples would require an additional normalization and validation step, which we leave to future work. Approximately half of the evaluation set (5,267 messages) contained a symptom event, with the organ-system role distributed across musculoskeletal, neurological, dermatological, and gastrointestinal categories. Medication events were present in 3,794 messages, of which approximately twelve percent mentioned a medication side-effect concern. Logistics events were present in approximately twenty-three percent of messages, covering policy inquiries, health IT concerns, and insurance questions, among others. Patient messages also carried urgent non-medical concerns, for example about financial status or living situation, at non-trivial frequency.

#### Human Evaluation of Event Detection and Role Extraction

We assessed extraction quality through manual review by two NLP researchers. For each event type, both annotators judged the same 30 random samples. For each sample, annotators decided (i) whether each detected event type was correctly identified (detection precision) and (ii) among correctly identified event types, whether the extracted arguments were correct (argument accuracy).

Table 5 reports per-event detection precision and argument accuracy, shown per annotator and averaged, with the number of items reviewed, 95% confidence intervals, and inter-annotator agreement. The framework produced accurate extractions for the majority of events in the random samples, because symptom, medication, and patient context reached a detection precision of 1.00 under both annotators, and medical needs and diagnostic test were at or above 0.94 on average. Argument accuracy was uniformly high across events (0.96–0.99). Inter-annotator agreement was high overall, 91% on the event-type verdict across 826 co-judged items (excluding treatment response, which was evaluated on a separate sample).

**Table 5.** Two-annotator human evaluation of the PortalEvent framework on 30 randomly selected samples. *Items* is the number of extracted event instances reviewed. *Detection precision* is the fraction of items whose event type was correctly identified, shown per annotator (A / B) and averaged, with a 95% Wilson confidence interval on the average. *Argument accuracy* is the fraction of correctly-typed items whose extracted argument was correct, averaged across annotators. *Argument Agreements* are inter-annotator percent agreement over co-judged items, with *n* shown. argument agreement is computed only over items both annotators judged correctly typed, so its *n* is smaller. *Treatment response was evaluated on a separate targeted sample of 30 messages.

| **Event** | **Items** | **Detection Precision** |  | **Argument Acc.** | **Agreement** |  |
|---|---|---|---|---|---|---|
|  |  | A / B | Avg [95% CI] | Avg | Event Type | Argument |
| Symptom | 252 | 1.00 / 1.00 | 1.00 [0.98–1.00] | 0.97 | 1.00 (*n*=252) | 0.95 (*n*=250) |
| Medication | 83 | 1.00 / 1.00 | 1.00 [0.96–1.00] | 0.96 | 1.00 (*n*=82) | 0.95 (*n*=82) |
| Patient Context | 110 | 1.00 / 1.00 | 1.00 [0.97–1.00] | 0.98 | 1.00 (*n*=110) | 0.95 (*n*=110) |
| Medical Needs | 175 | 0.94 / 1.00 | 0.97 [0.93–0.99] | 0.99 | 0.94 (*n*=174) | 0.98 (*n*=163) |
| Logistics | 80 | 0.62 / 0.88 | 0.75 [0.65–0.83] | 0.97 | 0.50 (*n*=80) | 0.95 (*n*=40) |
| Social | 80 | 0.80 / 1.00 | 0.90 [0.81–0.95] | 0.98 | 0.80 (*n*=80) | 0.97 (*n*=64) |
| Diagnostic Test | 48 | 0.88 / 1.00 | 0.94 [0.83–0.98] | 0.98 | 0.88 (*n*=48) | 0.95 (*n*=42) |
| *Targeted evaluation on a separate 30-message sample:* |  |  |  |  |  |  |
| Treatment Response | 119 | 0.53 / 1.00 | 0.77 [0.69–0.84] | 0.96 | 0.53 (*n*=118) | 0.92 (*n*=63) |

The lower averages for logistics (0.75) and social (0.90) were largely due to annotator disagreement: for logistics the two annotators reported 0.62 and 0.88, and for social 0.80 and 1.00, with corresponding type agreement of 0.50 and 0.80. These are cases where the boundary between event types was genuinely contested.

Because the original random sample contained no treatment response items, we evaluated this event separately on a targeted sample of 30 messages in which a treatment response event was detected, annotated by the same two annotators under the same protocol. Treatment response reached an argument accuracy of 0.96 and argument agreement of 0.92, but a lower average detection precision of 0.77 and type agreement of 0.53. This divergence is consistent with the failure modes discussed in the Limitations, where the LLM model tends to assign medication-related responses to the treatment response event, which is defined to be non-medication treatment.

### Ontology-guided Characterization of Clinician Follow-Up Questions

Figure 9 shows the distribution of clinician follow-up questions per message in this subset. The majority of messages received one clinician follow-up question (20,157), with another 6,935 receiving two and 2,287 receiving three. The distribution was right-skewed with a long tail. Using the standard interquartile range rule, Tukey’s rule, messages with four or more clinician follow-up questions were classified as outliers (above the 3.5 threshold), and a small number of responses contained as many as 21 follow-up questions.

**Figure 9.**
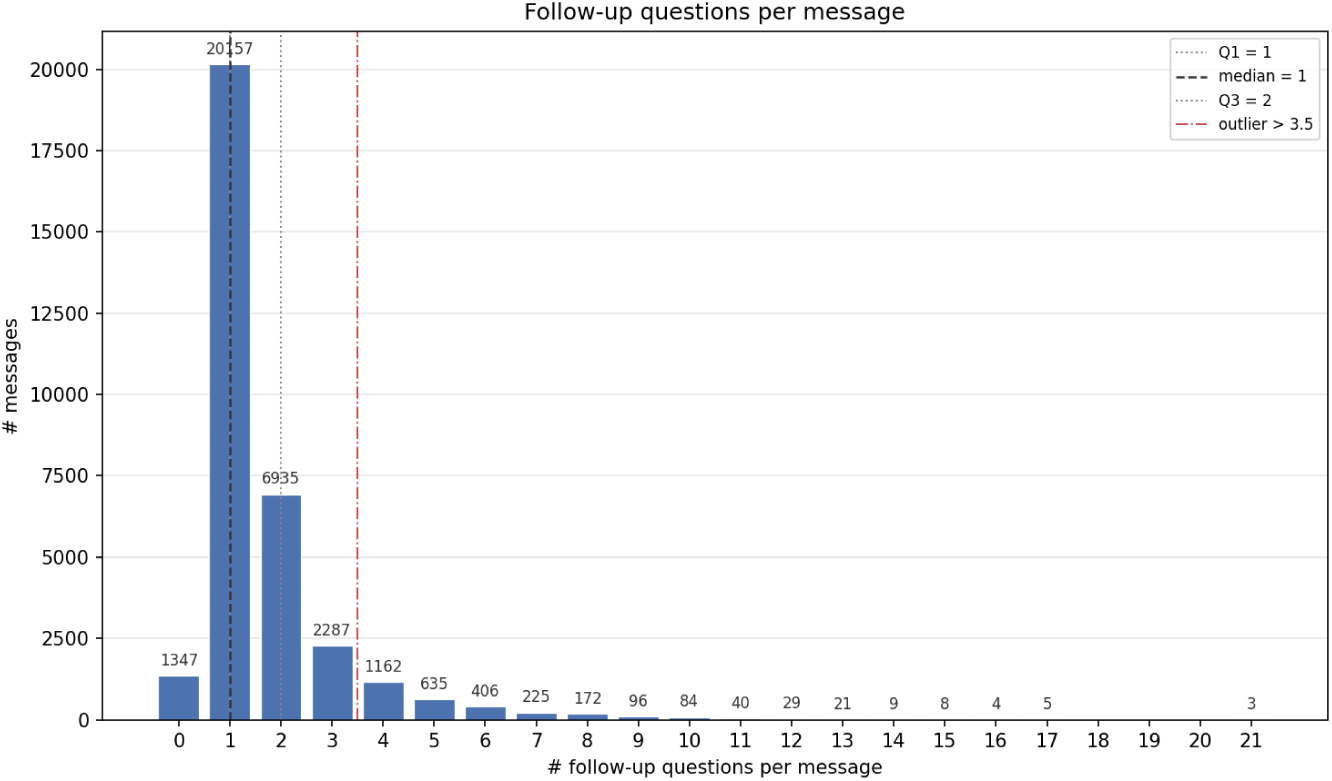
Distribution of follow-up questions per message in the subset. Most clinician responses contain one or two follow-up questions. Responses with four or more are classified as outliers relative to the interquartile range.

#### Event Types Targeted by Clinician Follow-Up Questions

Table 6 reports the number and percentage of follow-up questions for each event type, and the number of their corresponding messages. Symptom was the most targeted event type, accounting for 40.4% of all follow-up questions for 10,291 messages. Medication and medical needs were nearly tied by question share (20.2% each), but medical needs questions were in more messages (10,076 versus 8,965), indicating that medication tends to generate several questions within a message whereas medical needs are spread more evenly across messages. The remaining event types, logistics, patient context, diagnostic test, treatment response, and social, each accounted for under 11% of questions.

**Table 6.** Event types targeted by clinician follow-up questions. *Questions* is the number of follow-up questions targeting each event type and *% Q (n/N)* its share of all 57,321 follow-up questions with the underlying fraction shown. a question may target more than one event type. *Messages* is the number of distinct messages whose follow-up questions target that event type.

| **Event Type** | **Questions** | **% Q (n/N)** | **Messages** |
|---|---|---|---|
| Symptom | 23,163 | 40.4% (23,163/57,321) | 10,291 |
| Medication | 11,560 | 20.2% (11,560/57,321) | 8,965 |
| Medical Needs | 11,551 | 20.2% (11,551/57,321) | 10,076 |
| Logistics | 5,920 | 10.3% (5,920/57,321) | 5,274 |
| Patient Context | 3,703 | 6.5% (3,703/57,321) | 3,127 |
| Diagnostic Test | 2,616 | 4.6% (2,616/57,321) | 2,391 |
| Treatment Response | 2,522 | 4.4% (2,522/57,321) | 2,307 |
| Social | 1,578 | 2.8% (1,578/57,321) | 1,366 |

Two corpus-level patterns follow. First, medical needs was the second-most-frequent event type, indicating that requests such as scheduling, referrals, documentation content accounted for a share of clinician follow-up comparable to medication-related content. Second, when the event types were partitioned into clinical (symptom, medication, diagnostic test, patient context, treatment response) and administrative or social (medical needs, logistics, social), the clinical category accounted for 69.6%(43,564/62,613) event-type instances in the corpus and the administrative or social category accounted for 30.4%(19,049/62,613). The total exceeds the 57,321 questions because a single question may target more than one event type.

##### Validation of event-type LLM classification

The distributions in this section depend on the framework correctly identifying which event types and roles each follow-up question targets. To assess this, we sampled 200 follow-up questions and had a doctoral researcher working in medical NLP review each classification, annotating whether the assigned event type was correct (detection precision) and, among correctly-typed judgments, whether the assigned role was also correct (role accuracy). Because a single question may target more than one event type, the 200 questions yielded 248 event-type judgments and 202 role judgments. Event-type results are reported here, and role-level results in the Roles Targeted by Clinician Follow-Up Questions section.

Table 7 reports detection precision by event type. Overall detection precision was 0.87 (216 of 248 judgments), indicating that the event-type distribution reported above is largely reliable. Precision was highest for the event types that dominate the corpus, symptom (0.94) and medical needs (0.94), which together accounted for the majority of event-type judgments, along with diagnostic test (1.00) and logistics (0.90). Medication was the weakest of the high-volume event types at 0.79, and because it rests on 43 judgments this estimate is reasonably precise, marking medication classification as the clearest target for improvement.

**Table 7.** Human validation of event-type classification on 200 sampled follow-up questions. *Judgments* is the number of event-type judgments, which exceeds the number of questions because a single question may target multiple event types. *Detection precision* is the fraction of judgments in which the assigned event type was correct, with a 95% Wilson confidence interval. Overall figures are item-weighted across all judgments.

| **Event Type** | **Judgments** | **Detection Precision**<br>% (n/N) [95% CI] |
|---|---|---|
| Symptom | 90 | 94% (85/90) [87–97] |
| Medical Needs | 48 | 94% (45/48) [83–98] |
| Medication | 43 | 79% (34/43) [65–89] |
| Logistics | 20 | 90% (18/20) [70–97] |
| Diagnostic Test | 13 | 100% (13/13) [77–100] |
| Patient Context | 12 | 50% (6/12) [25–75] |
| Treatment Response | 12 | 50% (6/12) [25–75] |
| Social | 10 | 90% (9/10) [60–98] |
| **Overall** | **248** | **87% (216/248) [82–91]** |

Patient context and treatment response showed the lowest precision at 0.50, but both rest on only 12 judgments, giving confidence intervals that span 0.25 to 0.75. These figures should therefore be read as provisional rather than as evidence of a stable 50% error rate. Both are among the least frequently targeted event types in the corpus, so they contribute little to the aggregate distributions, which is also why the item-weighted overall precision of 0.87 exceeds the unweighted mean across event types of 0.81.

#### Roles Targeted by Clinician Follow-Up Questions

Within each event type, the share of questions targeting each role varies in shape. Some events have a single dominant role, while others have several roles at comparable frequency. Percentages below are taken relative to the number of questions targeting that event type, and may sum to more than 100% because a question may target multiple roles within an event type.

Within the symptom event (Table 8), the *symptoms* role dominates, targeted by 7,935 questions (34.3% of symptom-targeting questions) across 4,219 messages, more than twice the next most frequent role, *active* (3,044 questions, 2,675 messages). The five leading roles are *symptoms*, *active*, *symptom_location*, *progression_of_symptom*, and *aggravator*. The gap between the question and message counts for *symptoms* (7,935 questions but 4,219 messages) indicates that a single message often prompts more than one symptom question.

**Table 8.** Roles targeted by clinician follow-up questions within the symptom event. *Questions* is the number of questions targeting each role and *% Q (n/N)* its share of the 23,163 symptom-targeting questions with the underlying fraction shown.

| Role | Questions | % Q (n/N) | Messages |
|---|---|---|---|
| symptoms | 7,935 | 34.3% (7,935/23,163) | 4,219 |
| active | 3,044 | 13.1% (3,044/23,163) | 2,675 |
| symptom location | 2,201 | 9.5% (2,201/23,163) | 1,655 |
| progression of symptom | 2,044 | 8.8% (2,044/23,163) | 1,804 |
| aggravator | 1,895 | 8.2% (1,895/23,163) | 1,568 |
| self treatment | 1,617 | 7.0% (1,617/23,163) | 1,416 |
| organ system | 1,340 | 5.8% (1,340/23,163) | 1,114 |
| frequency | 1,251 | 5.4% (1,251/23,163) | 1,040 |
| onset | 949 | 4.1% (949/23,163) | 890 |
| functional limitation | 777 | 3.4% (777/23,163) | 704 |
| duration | 751 | 3.2% (751/23,163) | 713 |
| symptom severity | 598 | 2.6% (598/23,163) | 544 |
| symptom type | 441 | 1.9% (441/23,163) | 407 |
| pain quality | 431 | 1.9% (431/23,163) | 371 |
| acutely onset | 196 | 0.8% (196/23,163) | 191 |

Within the medication event (Table 9), the *medications* role, which captures the drug identity, is the most targeted (4,715 questions, 40.8%, across 3,956 messages), followed by *management* (3,078 questions, 26.6%). Together these two roles account for over two-thirds of medication-targeting questions, indicating that clinician follow-up on medications centers on establishing which drug is involved and how its use should be adjusted. The fulfillment- and effect-related roles, *prescription_request* (11.9%), *efficacy* (9.3%), *pharmacy* (8.9%), and *side_effects* (6.2%), form a middle tier, while *interaction_concern* is rarely targeted (172 questions, 1.5%).

**Table 9.**
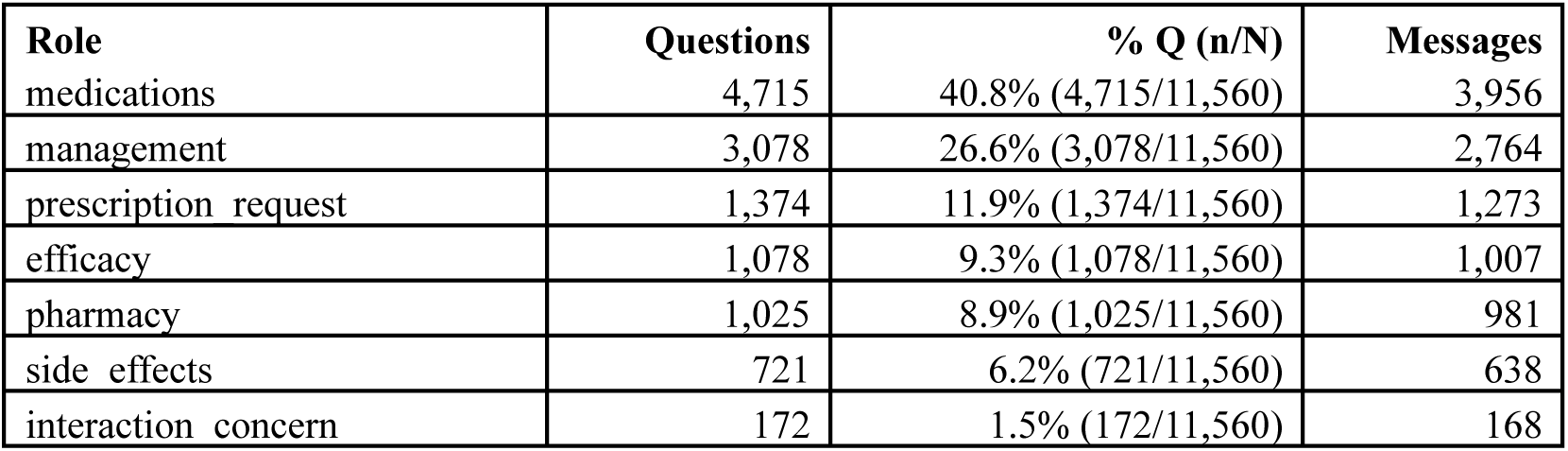
Roles targeted by clinician follow-up questions within the medication event. *Questions* is the number of questions targeting each role and *% Q (n/N)* its share of the 11,560 medication-targeting questions with the underlying fraction shown.

| Role | Questions | % Q (n/N) | Messages |
|---|---|---|---|
| medications | 4,715 | 40.8% (4,715/11,560) | 3,956 |
| management | 3,078 | 26.6% (3,078/11,560) | 2,764 |
| prescription_request | 1,374 | 11.9% (1,374/11,560) | 1,273 |
| efficacy | 1,078 | 9.3% (1,078/11,560) | 1,007 |
| pharmacy | 1,025 | 8.9% (1,025/11,560) | 981 |
| side_effects | 721 | 6.2% (721/11,560) | 638 |
| interaction_concern | 172 | 1.5% (172/11,560) | 168 |

Within the patient context event (Table 10), *medical_history* is the dominant role, targeted by 1,335 questions (36.1%) across 1,223 messages, well ahead of any other role. It is followed by *pregnancy_status* (426 questions, 11.5%), *medication_history* (388, 10.5%), and *emergency_event* (265, 7.2%). The remaining six roles each account for under 6% of patient-context-targeting questions.

**Table 10.**
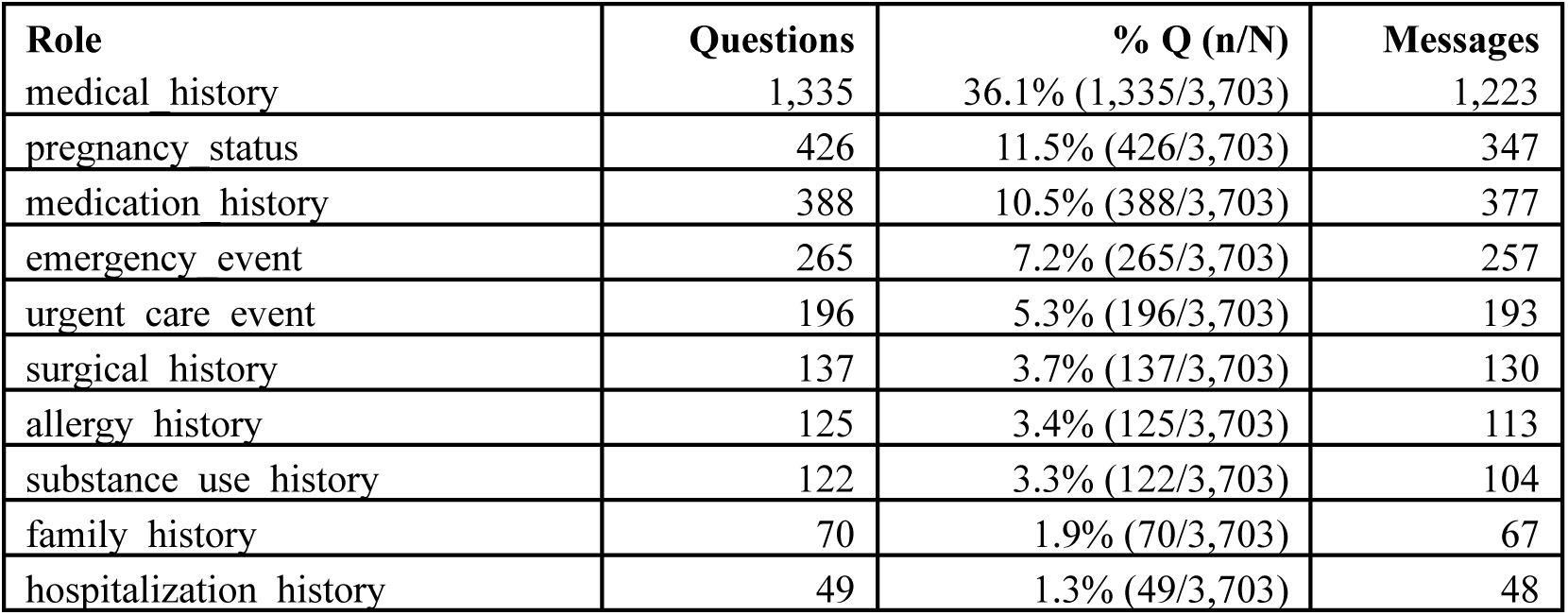
Roles targeted by clinician follow-up questions within the patient context event. *Questions* is the number of questions targeting each role and *% Q (n/N)* its share of the 3,703 patient-context-targeting questions with the underlying fraction shown.

Within the medical needs event (Table 11), the leading roles are *appointment_new* (3,848 questions, 33.3%), *appointment_time* (2,224, 19.3%), and *personnel_referrals* (1,819, 15.7%), which together account for over two-thirds of medical-needs-targeting questions. A second tier covers *appointment_type* (9.5%), *clinical_documentation* (7.9%), and *appointment_reschedule* (7.1%). The least frequent role is *appointment_cancellation* (123 questions, 1.1%).

**Table 11.** Roles targeted by clinician follow-up questions within the medical needs event. *Questions* is the number of questions targeting each role and *% Q (n/N)* its share of the 11,551 medical-needs-targeting questions with the underlying fraction shown.

| **Role** | **Questions** | **% Q (n/N)** | **Messages** |
|---|---|---|---|
| appointment_new | 3,848 | 33.3% (3,848/11,551) | 3,678 |
| appointment_time | 2,224 | 19.3% (2,224/11,551) | 2,145 |
| personnel_referrals | 1,819 | 15.7% (1,819/11,551) | 1,686 |
| appointment_type | 1,099 | 9.5% (1,099/11,551) | 1,073 |
| clinical_documentation | 918 | 7.9% (918/11,551) | 750 |
| appointment_reschedule | 821 | 7.1% (821/11,551) | 781 |
| follow_up | 647 | 5.6% (647/11,551) | 637 |
| appointment_confirmation | 559 | 4.8% (559/11,551) | 555 |
| medical_equipment | 377 | 3.3% (377/11,551) | 354 |
| medical_access_issue | 232 | 2.0% (232/11,551) | 229 |
| appointment_cancellation | 123 | 1.1% (123/11,551) | 122 |

Within the logistics event (Table 12), *facility_inquiry* (1,006 questions, 17.0%) and *contact_information_request* (930, 15.7%) are the leading roles, followed by *insurance_inquiry* (505, 8.5%) and *medical_records* (455, 7.7%). The least frequent role is *billing_inquiry* (93 questions, 1.6%).

**Table 12.** Roles targeted by clinician follow-up questions within the logistics event. *Questions* is the number of questions targeting each role and *% Q (n/N)* its share of the 5,920 logistics-targeting questions with the underlying fraction shown.

| **Role** | **Questions** | **% Q (n/N)** | **Messages** |
|---|---|---|---|
| facility_inquiry | 1,006 | 17.0% (1,006/5,920) | 977 |
| contact_information_request | 930 | 15.7% (930/5,920) | 907 |
| insurance_inquiry | 505 | 8.5% (505/5,920) | 458 |
| medical_records | 455 | 7.7% (455/5,920) | 433 |
| contact_information_update | 365 | 6.2% (365/5,920) | 358 |
| prior_authorization | 287 | 4.8% (287/5,920) | 277 |
| health_it_concern | 214 | 3.6% (214/5,920) | 208 |
| policy_inquiry | 202 | 3.4% (202/5,920) | 200 |
| transportation_need | 139 | 2.3% (139/5,920) | 138 |
| billing_inquiry | 93 | 1.6% (93/5,920) | 87 |

Within the social event (Table 13), *lifestyle_concern* (570 questions, 36.1%) and *financial_concern* (394, 25.0%) are the leading roles, followed by *living_situation_concern* (189, 12.0%) and *relationship_concern* (171, 10.8%). The least frequent roles are *food_insecurity* (37, 2.3%), *caregiver_burden* (34, 2.2%), and *complaints* (22, 1.4%).

**Table 13.**
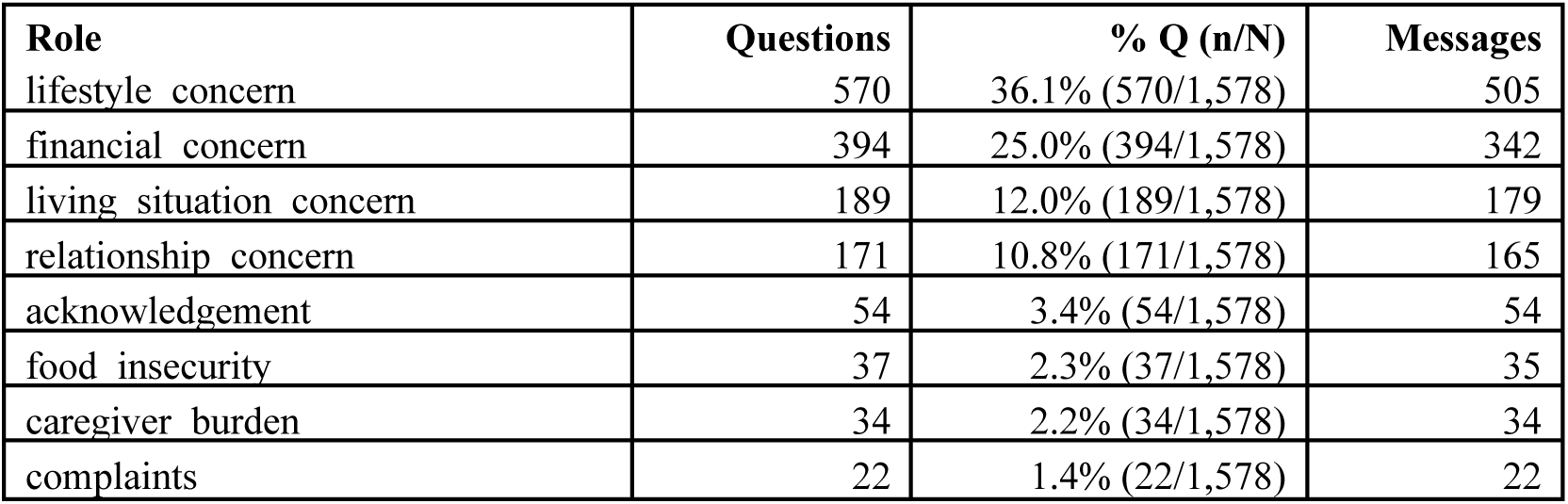
Roles targeted by clinician follow-up questions within the social event. *Questions* is the number of questions targeting each role and *% Q (n/N)* its share of the 1,578 social-targeting questions with the underlying fraction shown.

| Role | Questions | % Q (n/N) | Messages |
|---|---|---|---|
| lifestyle_concern | 570 | 36.1% (570/1,578) | 505 |
| financial_concern | 394 | 25.0% (394/1,578) | 342 |
| living_situation_concern | 189 | 12.0% (189/1,578) | 179 |
| relationship_concern | 171 | 10.8% (171/1,578) | 165 |
| acknowledgement | 54 | 3.4% (54/1,578) | 54 |
| food_insecurity | 37 | 2.3% (37/1,578) | 35 |
| caregiver_burden | 34 | 2.2% (34/1,578) | 34 |
| complaints | 22 | 1.4% (22/1,578) | 22 |

Within the diagnostic test event (Table 14), *test_scheduling* (1,000 questions, 38.2%) and *test_type* (963, 36.8%) are nearly tied as the leading roles, they account for three quarters of diagnostic-test-targeting questions. The result-related roles, *result_value* (8.6%), *result_interpretation* (5.4%), *interpretation_request* (4.4%), and *monitoring_trend* (3.1%), are each targeted much less often.

**Table 14.** Roles targeted by clinician follow-up questions within the diagnostic test event. *Questions* is the number of questions targeting each role and *% Q (n/N)* its share of the 2,616 diagnostic-test-targeting questions with the underlying fraction shown.

| **Role** | **Questions** | **% Q (n/N)** | **Messages** |
|---|---|---|---|
| test_scheduling | 1,000 | 38.2% (1,000/2,616) | 957 |
| test_type | 963 | 36.8% (963/2,616) | 897 |
| result_value | 225 | 8.6% (225/2,616) | 221 |
| result_interpretation | 142 | 5.4% (142/2,616) | 141 |
| interpretation_request | 114 | 4.4% (114/2,616) | 113 |
| monitoring_trend | 80 | 3.1% (80/2,616) | 78 |

Within the treatment response event (Table 15), *direction_of_response* (990 questions, 39.3%) and *treatment_reference* (895, 35.5%) are nearly tied and together account for the large majority of treatment-response-targeting questions.

**Table 15.** Roles targeted by clinician follow-up questions within the treatment response event. *Questions* is the number of questions targeting each role and *% Q (n/N)* its share of the 2,522 treatment-response-targeting questions with the underlying fraction shown.

| **Role** | **Questions** | **% Q (n/N)** | **Messages** |
|---|---|---|---|
| direction_of_response | 990 | 39.3% (990/2,522) | 945 |
| treatment_reference | 895 | 35.5% (895/2,522) | 849 |
| response | 89 | 3.5% (89/2,522) | 87 |
| comparison | 61 | 2.4% (61/2,522) | 60 |

##### Validation of role classification

Table 16 reports role accuracy on the same sample of 200 questions, computed over the 202 judgments in which the assigned event type was correct. Overall role accuracy was 0.89 (180 of 202), with six of the eight event types at or above 0.88. Symptom was the lowest of the high-volume event types at 0.86 (71 of 83), which may reflect that it has the largest role inventory of any event type, leaving more room for confusion between adjacent roles. Treatment response (0.80) and patient context (1.00) each rest on only 5 judgments, so their intervals are too wide to support conclusions.

**Table 16.** Human validation of role classification on the same 200 sampled follow-up questions. *Judgments* is the number of role judgments, counted only where the assigned event type was correct. *Role accuracy* is the fraction of these judgments in which the assigned role was also correct, with a 95% Wilson confidence interval. Overall figures are item-weighted across all judgments.

| **Event Type** | **Judgments** | **Role Accuracy**<br>% (n/N) [95% CI] |
|---|---|---|
| Symptom | 83 | 86% (71/83) [76–92] |
| Medical Needs | 43 | 93% (40/43) [81–98] |
| Medication | 34 | 91% (31/34) [77–97] |
| Logistics | 14 | 93% (13/14) [69–99] |
| Diagnostic Test | 10 | 90% (9/10) [60–98] |
| Patient Context | 5 | 100% (5/5) [57–100] |
| Treatment Response | 5 | 80% (4/5) [38–96] |
| Social | 8 | 88% (7/8) [53–98] |
| **Overall** | **202** | **89% (180/202) [84–93]** |

We note two limitations of this evaluation. Due to the cost of manual annotation, it was conducted by a single annotator, so we cannot report inter-annotator agreement as we do for the message-level evaluation. For the same reason as in event ontology extraction in messages, it measured precision rather than recall, assessing whether assigned event types and roles are correct rather than whether the framework missed content a question also targeted.

## Discussion

We introduced PortalEvent, an event ontology of 8 event types and 71 roles for patient portal communication, and we also designed a training-free extraction framework deployable on open-source LLMs. Event detection rates were broadly consistent across three open-source models spanning general-purpose and medical-domain families. Human evaluation confirmed high extraction quality, with strong detection precision for most event types and high inter-annotator agreement. The corpus-level analysis further showed that patient messages were frequently multi-topic, with administrative events such as medical needs co-occurring with clinical events such as symptom and medication.

### Event Detection and Role Extraction

#### Principal Findings

Event detection rates were broadly consistent across the three extraction models, with the relative ordering of common event types preserved. This indicates that the ontology’s event structure can be recoverable independently of the specific extractor. Medical needs and symptom events were the most frequent, each detected in over half of messages, while treatment response was the least frequent at 10.9 percent. The co-occurrence analysis showed that medical needs participated in three of the six strongest co-occurring pairs, including the two strongest, while logistics co-occurred with clinical events at below 10 percent. Human evaluation confirmed fairly high extraction quality, with detection precision at or above 0.90 for most event types and high inter-annotator agreement.

#### Implications

The high detection rates for medical needs and symptom events suggest that portal messaging is used as both a clinical and administrative channel. More than half of patient messages contained an administrative request such as scheduling, referral, or documentation, and a comparable proportion reported clinical symptoms. The intermediate rates for medication, patient context, social, and diagnostic test events indicate that these categories appear with moderate but not majority frequency, while the lower rates for logistics and treatment response correspond to their narrower scopes.

These rates have practical implications for downstream systems that operate on portal messages. The uneven distribution means that a small number of event types dominate message content, so a follow-up question generator that targets the most frequently sought roles within symptom and medical needs, could in principle cover a large share of the majority of cases a clinician would actually raise, while rarer event types can be handled by fallback behavior. The low rate of treatment response indicates that systems targeting it may see fewer training examples in a typical corpus, and the corresponding results should be interpreted accordingly.

The co-occurrence structure has implications for how portal messages are triaged. Administrative and clinical content appeared together in a message quite often, with medical needs co-occurring with symptom and social events in over a quarter of messages. Sometimes, co-occurrence of multiple event types in a message can occur from message exchanges between the patient and clinician. For instance, a message that begins with a single event, such as scheduling or medical needs, may lead to introducing more event types such as symptoms or medications in subsequent patient messages, depending on the interaction with the clinician. Logistics, by contrast, co-occurred with symptom, medication, and patient context at below 10 percent, which suggests that purely administrative inquiries such as insurance, billing, or records requests are largely self-contained and could be routed independently of messages requiring clinical interpretation. Logistics events appeared in roughly a quarter of messages, a volume that suggests potential value in automated support for tasks such as troubleshooting, document retrieval, and profile management. Patient messages also carried urgent non-medical concerns such as financial or living situations at non-trivial frequency. Prior work on portal-message triage has focused on medical urgency, these results suggest that AI-mediated triage solutions could be broadened to account for non-medical urgent content.

### Ontology-guided Characterization of Clinician Follow-Up Questions

#### Principal Findings

Clinician follow-up questions concentrated on a small set of event types. Within some event types, the questions followed a small set of event roles. The role that identifies the basic instance of an event, such as the *symptoms* role for *symptom reporting* event, the *medications* role for the *medication* event, *test scheduling* for *diagnostic test* event, appointment new for medical needs, and medical history for patient context, consistently ranked among the most frequently sought roles in its type. At the event-type level, the symptom type was targeted most often, followed by medical needs and medication at similar volume. Several roles defined in the ontology, such as interaction concern, appointment cancellation, billing inquiry, and comparison, were sought at low frequencies relative to the volume of questions targeting their event type. These findings reflect only follow-up questions asked through the patient portal. About 32,000 of the 146,000 message threads contained written follow-up questions, but clinicians also follow up by phone, which is common in portal communication flows. The portal-based counts therefore likely underestimate how often clinicians need additional information from patients.

#### Implications

The concentration of questions on the role that identifies the basic instance of each event suggests that a generation system could use these high-frequency roles as candidate targets, a possibility we leave to future work. More broadly, a clinician asks a follow-up question when a message lacks presence or clarity of certain information needed to act, the roles most frequently sought serve as a proxy for the information patients most often leave under-specified. Across event types, the clinician-sought roles fall into two recurring kinds: the identity of the entity in question, such as *which symptoms are present*, *which medications are involved*, *which test is meant*, or *the relevant medical history*, and the details needed to act on it, such as *appointment timing*, *test scheduling*, or *the location and progression of a symptom*. This indicates that patients tend to report that a concern exists without supplying the specifics a clinician needs, and a generation system grounded in these patterns can anticipate the particular information a message is likely to be missing rather than asking generic clarifying questions.

The event-type distribution indicates that follow-up is not predominantly clinical, because medical needs was the second most frequent target and scheduling, referral, and care-coordination content accounted for a share of clinician follow-up comparable to medication-related content. A follow-up generation system focused only on clinical content would therefore miss a large fraction of the questions clinicians actually ask. Some low-frequency roles in follow-up questions contribute more to characterizing the content of patient messages, indicating that the ontology captures message content beyond what is reflected in questioning behavior. Other low-frequency social roles, such as food insecurity and caregiver burden, are recognized social determinants of health that are rarely surfaced through asynchronous messaging, suggesting that targeted screening rather than spontaneous disclosure may be needed to capture them.

### Comparison with Prior Work

Prior characterizations of patient portal messages have largely operated at the level of message topics or categories [6–8], which supports routing but not fine-grained analysis about message content. More recent work has applied topic modeling and LLMs to surface themes in message corpora [10], and a related line extracts clinical entities from clinical text [32], but neither produces a structured, event-and-role representation of what a message contains. Our ontology instead specifies 71 roles across 8 event types, which allows extraction at a finer granularity than topic categorization while remaining interpretable at the level of individual roles.

Our construction and extraction methodology connects to a growing body of work that uses LLMs to build and populate ontologies [33,34,36,38]. This work has been applied mainly to knowledge-graph construction in general or technical domains, and its clinical instances target formal disease knowledge, entities and relations drawn from medical literature and clinical reports [41,42]. We adapt the same schema-guided, training-free paradigm to a different setting, patient-authored portal messages, and to a different unit of analysis, communicative events and their roles. Consistent with prior findings that general-purpose LLMs are effective zero- and few-shot clinical extractors [17], our framework requires no site-specific training data and can be applied to a private corpus without shared training data. Because message-event distributions may vary across patient populations and healthcare systems, applying the framework at a new site would require re-estimating these distributions locally.

### Limitations

The PortalEvent ontology does not adopt certain conventions of established event ontologies [15], such as predefined valid entity types per role. Future work could explore an entity-typed extension of the ontology. We note, however, that these conventions are less central to the present framework, which prioritizes argument-level normalization through discretization of the output space.

Because manual annotation by qualified reviewers is costly, human evaluation was conducted on a relatively small sample and focused on detection precision. For most event types the evidence at this scale was consistent, with detection precision at or above 0.94 and inter-annotator type agreement at or above 0.88 for five of the eight events, so a larger sample would be unlikely to overturn the conclusion that extraction quality is high for those types. Two events are exceptions. Logistics and treatment response showed both lower precision, 0.75 and 0.77, and substantially lower type agreement, 0.50 and 0.53, indicating that the annotators disagreed about what counts as a correct assignment rather than converging on a stable estimate. For these two events the reported figures should be treated as provisional. Argument accuracy was uniformly high across events, but role-level validation remains limited, and a broader role-level audit is left to future work.

Several further limitations constrain what our results establish. The corpus comes from a single health system, so the reported event and role distributions characterize this setting rather than portal messaging in general, and applying the framework at another site would require re-estimating them locally. The ontology was refined by a small expert panel, and a larger or differently composed panel might have drawn different role boundaries. Making the ontology publicly available allows other health systems to apply the framework to their own messages and report how well it generalizes, including event types or role arguments not captured in our setting. For example, a health system in a different region may have institution-specific events, such as local programs or administrative processes, that are uncommon in our data. Our annotation protocol measured precision but not recall, asking whether detected events and role arguments are correct rather than whether the framework missed content that was present, so the prevalence estimates we report are model-generated rather than validated against an exhaustively annotated reference. Finally, we do not evaluate any downstream system. Message completeness scoring, follow-up question generation, and triage are applications the representation is intended to support, but whether it improves them is an empirical question we do not answer here. Our follow-up question analysis includes only questions written in portal responses. Clinicians often follow up by phone, and although our data partially record whether a call took place, they do not capture whether or how many questions were asked during the call. To ensure data reliability, we exclude phone follow-up from this analysis, so the reported counts likely underestimate clinicians’ total follow-up questions.

The ontology is also, by design, fitted to the head of the distribution. Roles were retained when they recurred across the corpus, so the schema models common content well but does not cover the long tail of message and question types. Our extraction of follow-up questions reflects the same bias. Among candidate responses containing a question mark, the model did not identify an extractable follow-up question in roughly four percent, typically where the question was rhetorical or hedged rather than a substantive request, or where it was too specific to the message context to map onto a recurring role. Extending the ontology to cover tail phenomena, and characterizing what share of clinician questions they represent, remains open.

Although the LLM models make it possible to apply the ontology at scale, their extractions are not error-free. In our data, the models were not able to detect any events in 2.5% to 4.5% of messages. In the reviewed cases, these misses typically involved short messages, or messages that contained a valid event type the model failed to detect, which suggests limitations of the LLM. Errors also varied by event type. The treatment response event was the most difficult to extract. Two recurring failure modes were observed, extraction of medication-related responses under the treatment response event, and correct event assignment with an incorrect treatment argument. When applying the framework to a new dataset, users should review messages with missing events and event types with low precision to identify the causes of extraction errors.

### Conclusions

PortalEvent is a scalable and reproducible event ontology for patient portal messages, developed and validated in collaboration with three clinicians. We characterized both patient messages and clinician follow-up questions, providing a shared structured representation of what patients communicate and what clinicians ask about. This representation is intended to support applications such as message completeness scoring, follow-up question generation, and triage. The framework operates training-free and uses prompts that contain no protected health information, so that the prompts themselves can be shared across institutions. We do not evaluate these applications here, and doing so is left to future work.

## Supporting information

Multimedia Appendix 1: Event ontology definitions

Multimedia Appendix 2: Supplementary results

## Data Availability

The patient portal message corpus analyzed in this study contains sensitive patient information and cannot be publicly shared. The PortalEvent ontology specification is provided in Multimedia Appendix 1. The extraction prompts are available from the corresponding author on reasonable request.

## Authors’ Contributions

JY: Conceptualization, Formal analysis, Investigation, Methodology (follow-up question analysis), Software (follow-up question extraction), Validation (annotation), Visualization, Writing – original draft, Writing – review & editing. JG: Conceptualization, Formal analysis, Investigation, Validation, Methodology, Software, Visualization, Writing – review & editing. RR: Investigation, Validation (annotation), Writing – review & editing. PS: Validation, Writing – review & editing. TEB: Conceptualization (clinical domain expertise), Validation (ontology schema review and coverage annotation), Writing – review & editing. MK: Validation (ontology schema review), Writing – review & editing. SKL: Validation (ontology schema review), Writing – review & editing. SMP: Supervision, Writing – review & editing.

## Abbreviations

AI: artificial intelligence
DHMC: Dartmouth Hitchcock Medical Center
EAE: event argument extraction
EE: event extraction
EHR: electronic health record
GPU: graphics processing unit
HIPAA: Health Insurance Portability and Accountability Act
IQR: interquartile range
LLM: large language model
NLP: natural language processing
UMLS: Unified Medical Language System

