## Supplementary material for "PortalEvent: An Event-Based Ontology for Patient-Initiated Portal Communication": Multimedia Appendix 1: Event ontology definitions

### Event Ontology Definitions

The complete set of event types and roles in the PortalEvent ontology, grouped by event type, are listed in Tables S1 and S2.

Table S1. Event types in the PortalEvent ontology and their definitions.

| Event Type | # Roles | Definition |
|---|---|---|
| Symptom | 15 | The patient describes a physical, mental, or functional symptom they are experiencing or have experienced, together with details such as onset, duration, frequency, location, progression, intensity/severity, aggravating factors, quality, functional impact, etc. A symptom may span more than one organ system or symptom type. |
| Medication | 7 | The patient discusses one or more medications they are taking, have taken, or have been prescribed, together with details such as drug identity, dosage, regimen, prescription and refill requests, side effects, adherence, pharmacy, perceived efficacy, etc. |
| Patient Context | 10 | The patient provides background medical information about themselves, including past medical conditions, prior surgeries or hospitalizations, allergies, immunization history, family history, pregnancy status, substance-use history, and recent emergency or urgent-care visits, etc. |
| Medical Needs | 11 | The patient expresses a care-related need or request, including scheduling, rescheduling, cancelling, or confirming an appointment, referrals to personnel in primary care or other specialties, medical equipment, follow-up on a care plan or on recently performed tests or studies, access barriers, and clinical documentation requests, etc. |
| Logistics | 10 | The patient raises an administrative or operational matter relating to the practice or the health system, including insurance, billing, medical records, contact-information updates, prior authorization, transportation, facility hours or policies, health-IT concerns, etc. |
| Social | 8 | The patient describes non-medical life circumstances that may bear on their health or care, including employment and financial hardship, housing, lifestyle, caregiver burden, food access, interpersonal relationships, and non-medical complaints such as dissatisfaction with a provider or reported inaccuracies in their visit notes or records, etc. |
| Diagnostic Test | 6 | The patient discusses a diagnostic test or imaging study, including scheduling, the type of test, reported result values, the patient's interpretation of the result, requests for help interpreting it, trends across repeated tests, etc. |

| Event Type | # Roles | Definition |
|---|---|---|
| Treatment Response | 4 | The patient reports the outcome of a non-medication treatment (procedure, therapy, surgery, or similar), including the treatment in question, the direction of response, comparison to prior treatments, and the response or reaction attributed to it. |

Table S2. Full set of roles in the PortalEvent ontology, grouped by event type. The Type column indicates each role's output space: BIN (binary), CAT (closed categorical), FREE (free text), or MULTI (multiple values).

| Role | Type | Definition |
|---|---|---|
| **Symptom Event** | | |
| `active` | CAT | Whether the symptoms are currently active, negated, or resolved. |
| `acutely_onset` | CAT | Whether the symptoms are acute or chronic in nature. |
| `aggravator` | FREE | Patient-attributed factor that triggers or worsens the symptom, such as precipitating activities, exposures, positions, and contextual factors. |
| `self_treatment` | CAT | Type of self-treatment attempted (OTC medication, herbal remedy, home physical therapy, rest and lifestyle, home monitoring, other, none). |
| `organ_system` | MULTI | Organ system or systems associated with the symptoms (cardiovascular, respiratory, gastrointestinal, neurological, musculoskeletal, dermatological, endocrine, genitourinary, ophthalmologic, ENT, hematologic, immunologic, psychiatric, reproductive, other). A symptom may implicate more than one system. |
| `symptom_type` | MULTI | Type or types of the symptom (physiological, mental/behavioral, cognitive, functional). A symptom may span more than one type. |
| `progression_of_symptom` | CAT | Whether the symptoms have gotten better, worse, or stayed the same. |
| `onset` | CAT | When the symptoms started (today, yesterday, past 7 days, past month, past year, over a year ago, none). |
| `duration` | FREE | How long each episode of the symptom lasts. Distinct from onset: captures per-episode duration, not how long the condition has been present. |
| `frequency` | CAT | How frequently the symptoms occur (constantly, hourly or more, daily, weekly, monthly, occasionally, intermittently). |
| `symptom_severity` | FREE | How severe the patient reports the symptom to be, whether stated qualitatively (mild, moderate, severe, unbearable) or on a numeric scale. |
| `symptoms` | MULTI | Comma-separated list of all symptoms mentioned by the patient. |
| `symptom_location` | FREE | Specific anatomical location or laterality of the symptom (e.g., left knee, right temple). Use `organ_system` for system-level classification. |

| Role | Type | Definition |
|---|---|---|
| functional_limitation | BIN | Whether the patient describes activities they can no longer perform due to symptoms. |
| pain_quality | FREE | Descriptive quality of pain (burning, sharp, dull, throbbing, pressure). |
| **Medication Event** | | |
| medications | FREE | For each medication mentioned, a tuple (name, dosage, routine): dosage as stated (e.g., 500mg) or none, routine as stated in the patient's own words (e.g., "twice a day", "as needed", "as prescribed") or none. |
| side_effects | BIN | Whether the patient reports side-effects or adverse reactions attributed to the medication. |
| prescription_request | BIN | Whether the patient requests a prescription, either a refill of an existing medication or a new prescription. |
| management | BIN | Whether the patient discusses strategy updates (dose changes, switching, samples, prescription changes). |
| interaction_concern | BIN | Whether the patient raises concerns about interactions with other medications or conditions. |
| pharmacy | FREE | The name or location of a pharmacy the patient mentions. |
| efficacy | CAT | Whether the patient reports medication(s) as effective, partially effective, ineffective, or mixed (contrasting outcomes across multiple medications). |
| **Patient Context** | | |
| pregnancy_status | BIN | Whether the patient is or may be pregnant. |
| medical_history | MULTI | Relevant past medical information provided by the patient, including past medical conditions and immunization history. |
| medication_history | MULTI | Current and prior medications the patient reports. |
| family_history | MULTI | Relevant family medical history provided by the patient. |
| emergency_event | BIN | Whether the patient reports a recent emergency healthcare event. |
| urgent_care_event | BIN | Whether the patient reports a recent urgent care visit. |
| allergy_history | CAT | Type of allergies reported (drug, food, environmental, multiple, none). |
| surgical_history | CAT | Type of past surgeries reported (orthopedic, abdominal, cardiac, reproductive, other, multiple, none). |
| hospitalization_history | CAT | Type of past hospitalizations reported (medical, surgical, psychiatric, obstetric, multiple, none). |
| substance_use_history | BIN | Whether the patient mentions any history of substance use (smoking, alcohol, drugs, vaping). |
| **Medical Needs** | | |
| appointment_new | BIN | Whether the patient requests to schedule a new appointment. |

| Role | Type | Definition |
|---|---|---|
| appointment_reschedule | BIN | Whether the patient requests to reschedule an existing appointment. |
| appointment_cancellation | BIN | Whether the patient requests to cancel an existing appointment. |
| appointment_confirmation | BIN | Whether the patient confirms or acknowledges an upcoming appointment. |
| medical_equipment | BIN | Requests or discussions about medical equipment. |
| personnel_referrals | BIN | Requests or discussions about medical personnel or referrals for ongoing care, in primary care or other specialties (cardiology, endocrinology). |
| follow_up | BIN | Discussion about care plan follow-up without a new or worsening problem, including follow-up on tests or studies recently performed. |
| medical_access_issue | CAT | Type of barrier the patient describes to accessing medical care (insurance, cost, provider availability, transportation, multiple, other, none). |
| clinical_documentation | BIN | Whether the patient requests clinical documentation (FMLA forms, school notes, disability letters, medical summaries). |
| appointment_time | FREE | The requested or scheduled appointment time or time preference mentioned by the patient. |
| appointment_type | CAT | Whether the patient specifies an appointment modality (in-person, telehealth, or none). |
| **Logistics** | | |
| contact_information_request | BIN | Whether the patient requests contact information for a provider or facility. |
| contact_information_update | BIN | Whether the patient provides or updates their own contact information. |
| facility_inquiry | BIN | Whether the patient asks about facility location, hours, or directions. |
| policy_inquiry | BIN | Whether the patient asks about facility policies or procedures. |
| insurance_inquiry | BIN | Whether the patient mentions or asks about insurance coverage or authorization. |
| billing_inquiry | BIN | Whether the patient mentions or asks about billing, charges, or payments. |
| medical_records | CAT | Whether the patient discusses their medical records (request copy, request correction, status inquiry, or none). |
| health_it_concern | BIN | Whether the patient has a concern about health IT (portal, EHR, telehealth). |
| transportation_need | BIN | Whether the patient expresses a transportation need or concern. |
| prior_authorization | BIN | Whether the patient mentions or asks about prior authorization status for a medication, test, or procedure. |
| **Social** | | |
| acknowledgement | BIN | Whether the text contains expressions of gratitude, satisfaction, or agreement. |

| Role | Type | Definition |
| --- | --- | --- |
| complaints | BIN | Whether the patient complains about a non-medical issue, including dissatisfaction with their current provider or care experience, and reported inaccuracies in released clinical information such as progress notes or visit summaries. |
| relationship_concern | BIN | Whether the patient describes non-medical interpersonal concerns. |
| living_situation_concern | BIN | Whether the patient describes concerns about their living or housing situation (arrangements, instability, unsafe conditions, relocation, eviction). |
| financial_concern | BIN | Whether the patient describes employment and financial hardship concerns. |
| lifestyle_concern | BIN | Whether the patient describes concerns about lifestyle or daily functioning (mobility, memory, self-agency, well-being). |
| food_insecurity | BIN | Whether the patient reports difficulty affording or accessing food. |
| caregiver_burden | BIN | Whether the patient describes burden, strain, or exhaustion from caregiving responsibilities. |
| **Diagnostic Test** | | |
| test_scheduling | BIN | Whether the patient requests or discusses scheduling a test. |
| test_type | FREE | The type of test or imaging the patient discusses (lab, imaging, biopsy, screening). |
| result_value | FREE | The test result value or finding as reported by the patient. |
| result_interpretation | CAT | Whether the patient describes the result as normal, abnormal, critical, unclear, or none when no result is described. |
| interpretation_request | BIN | Whether the patient asks for help understanding a test result. |
| monitoring_trend | CAT | Whether the patient describes a trend in repeated test results (improving, worsening, stable). |
| **Treatment Response** | | |
| treatment_reference | FREE | The non-medication treatment (procedure, therapy, surgery, etc.) the patient attributes the response to. |
| direction_of_response | CAT | Whether the patient reports a non-medication treatment as effective, partially effective, ineffective, worsening, or none when no non-medication treatment response is described. |
| comparison | BIN | Whether the patient compares a non-medication treatment to a prior treatment or no treatment. |
| response | BIN | Whether the patient reports any side-effects or adverse reactions they attribute to a non-medication treatment. |
