## Supplementary material for "PortalEvent: An Event-Based Ontology for Patient-Initiated Portal Communication": Multimedia Appendix 2: Supplementary results

### Results

Table S3. Corpus-level characterization produced by the PortalEvent framework (GPT-OSS-20B) on 10,000 randomly sampled patient-initiated messages. For each event, *n* is the number of messages in which the event was detected. For most roles, the percentage is the share of those n messages in which the role was instantiated. For roles marked with an asterisk (*), percentages show the distribution of values among messages in which that role was instantiated. Roles with a categorical output space are broken out by value, roles with a binary output space are marked with "—" in the value column.

| **Event** | ***n*** | **Role** | **Value** | **%** |
|---|---|---|---|---|
| Symptom | 5,267 | Organ System* | Musculoskeletal | 26.5 |
| | | | Neurological | 20.6 |
| | | | Dermatological | 16.2 |
| | | | Gastrointestinal | 15.9 |
| | | | Respiratory | 10.6 |
| | | | Psychiatric | 10.2 |
| | | Symptom Type* | Physiological | 85.4 |
| | | | Functional | 6.9 |
| | | | Mental/Behavioral | 6.1 |
| | | | Cognitive | 1.6 |
| Medication | 3,794 | Management | — | 52.3 |
| | | Prescription Request | — | 15.0 |
| | | Side Effects | — | 12.6 |
| | | Interaction Concern | — | 10.0 |
| Patient Context | 4,026 | Pregnancy Status | — | 6.1 |
| | | Emergency Event | — | 5.6 |
| | | Urgent Care Event | — | 1.2 |
| | | Medical History (top 5)* | Diagnoses | 25.4 |
| | | | Chronic conditions | 24.8 |
| | | | Known health conditions | 23.6 |
| | | | Prior surgeries | 11.9 |
| | | | Hospitalizations | 2.0 |
| Medical Needs | 5,534 | Appointment Confirmation | — | 19.9 |
| | | Appointment New | — | 18.8 |
| | | Appointment Reschedule | — | 4.5 |
| | | Appointment Cancellation | — | 0.9 |
| | | Appointment Type* | in-person | 63.0 |
| | | | telehealth | 37.0 |

| **Event** | ***n*** | **Role** | **Value** | **%** |
|---|---|---|---|---|
| Logistics | 2,329 | Policy Inquiry | — | 25.8 |
| | | Health IT Concern | — | 18.9 |
| | | Insurance Inquiry | — | 16.5 |
| | | Transportation Need | — | 9.5 |
| | | Prior Authorization | — | 6.7 |
| | | Billing Inquiry | — | 6.6 |
| | | Contact Information Request | — | 5.5 |
| | | Facility Inquiry | — | 4.1 |
| | | Contact Information Update | — | 0.7 |
| Social | 4,035 | Lifestyle Concern | — | 56.1 |
| | | Complaints | — | 39.5 |
| | | Acknowledgement | — | 19.6 |
| | | Financial Concern | — | 9.5 |
| | | Relationship Concern | — | 6.7 |
| | | Living Situation Concern | — | 1.8 |
| | | Caregiver Burden | — | 1.5 |
| | | Food Insecurity | — | 0.1 |
| Diagnostic Test | 2,972 | Test Scheduling | — | 51.4 |
| Treatment Response | 1,091 | Response | — | 35.4 |

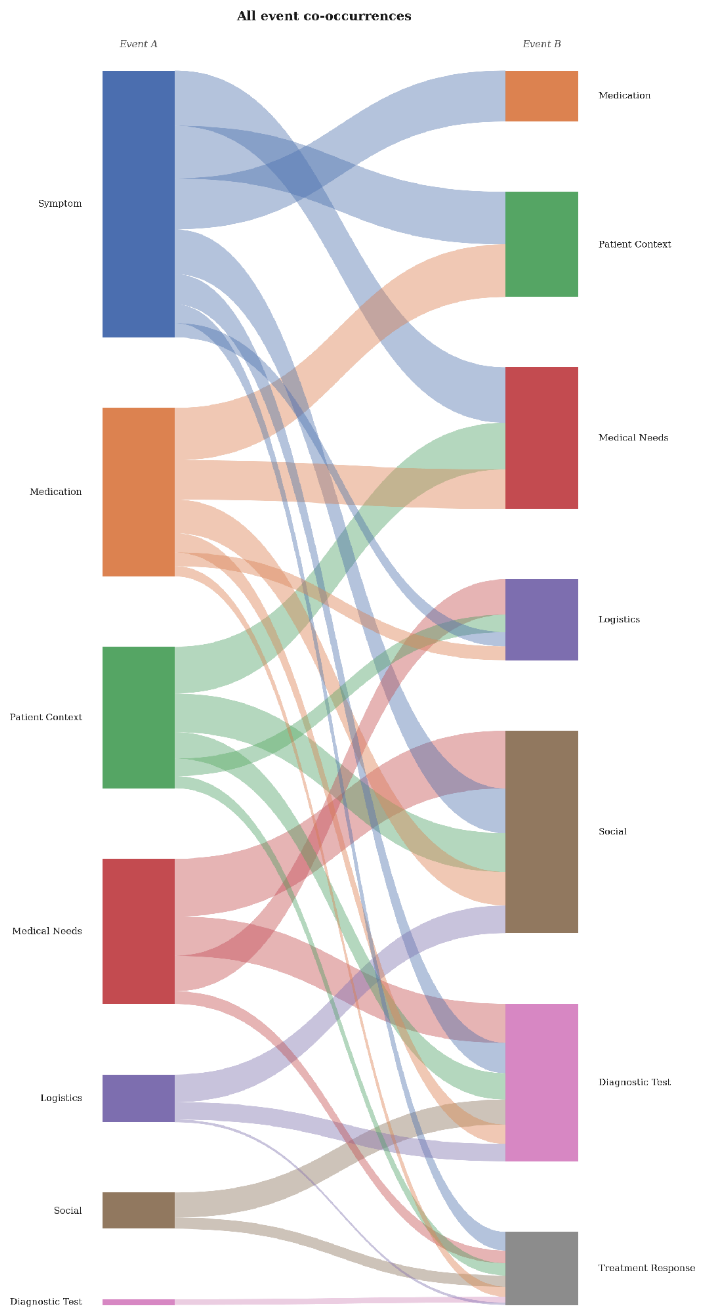


Figure S1. All pairwise event co-occurrences in the 10,000-message sample, shown as a Sankey diagram. Co-occurrence is symmetric; the left (Event A) and right (Event B) columns are a layout device and do not indicate direction. Each ribbon connects a pair of event types, with width proportional to the percentage of messages in which both events were detected. Ribbons are colored by the left-hand event.
